# Machine learning identifies clinical sepsis phenotypes that translate to the plasma proteome: a prospective cohort study

**DOI:** 10.1101/2025.04.11.25325574

**Authors:** Thilo Bracht, Maike Weber, Kerstin Kappler, Lars Palmowski, Malte Bayer, Karin Schork, Tim Rahmel, Matthias Unterberg, Helge Haberl, Alexander Wolf, Björn Koos, Katharina Rump, Dominik Ziehe, Ulrich Limper, Dietrich Henzler, Stefan Felix Ehrentraut, Thilo von Groote, Alexander Zarbock, Martin Eisenacher, Michael Adamzik, Barbara Sitek, Hartmuth Nowak

**Affiliations:** Department of Anesthesiology, Intensive Care Medicine and Pain Therapy, University Hospital Knappschaftskrankenhaus Bochum, Bochum, Germany; Medizinisches Proteom-Center, Ruhr-University Bochum, Bochum, Germany; Center for Protein Diagnostics (PRODI), Medical Proteome Analysis, Ruhr-University Bochum, Bochum, Germany; CUBiMed.RUB, Core Unit Bioinformatics, Medical Faculty, Ruhr University Bochum, Bochum, Germany; Center for Artificial Intelligence, Medical Informatics and Data Science, University Hospital Knappschaftskrankenhaus Bochum, Bochum, Germany; Department of Anesthesiology and Operative Intensive Care Medicine, University of Witten/Herdecke, Cologne Merheim Medical School, 51109 Cologne, Germany; Department of Anesthesiology, Surgical Intensive Care, Emergency and Pain Medicine, Ruhr-University Bochum, Klinikum Herford, 32049 Herford, Germany; Klinik für Anästhesiologie und Operative Intensivmedizin, Universitätsklinikum Bonn, 53127 Bonn, Germany; Department of Anesthesiology, Intensive Care and Pain Medicine, University Hospital of Münster, Münster, Germany

**Keywords:** Artificial Intelligence, Proteomics, Cluster Analysis, Precision Medicine, Mass Spectrometry

## Abstract

**Background:** Sepsis therapy is still limited to treatment of the underlying infection and supportive measures. To date, various sepsis subtypes were proposed, but therapeutic options addressing the molecular changes of sepsis were not identified. With the aim of a future individualized therapy, we used machine learning (ML) to identify clinical phenotypes and their temporal development in a prospective, multicenter sepsis cohort and characterized them using plasma proteomics.

**Methods:** Routine clinical data and blood samples were collected from 384 patients. Sepsis phenotypes were identified based on clinical measurements and plasma samples from 301 patients were analyzed using mass spectrometry. The obtained data were evaluated in relation to the phenotypes, and supervised ML models were developed enabling prospective phenotype classification and determination of key features distinguishing the phenotypes.

**Results:** Three phenotypes and their progression across four time points in sepsis were identified. Cluster C was characterized by the highest disease severity and multi-organ failure with leading liver failure. Cluster B showed relevant organ failure, with renal damage being particularly prominent in comparison to cluster A. Time course analysis showed a strong association of cluster C with mortality and dynamic properties of cluster B. The plasma proteome reflected the clinical features of the phenotypes and revealed excessive consumption of complement and coagulation factors in severe sepsis. Supervised ML models allow the assignment of patients based on only seven widely available features.

**Conclusions:** The identified clinical phenotypes reflected varying degrees of sepsis severity and were mirrored in the plasma proteome. Proteomic profiling offered novel insights into the molecular mechanisms underlying sepsis and enabled a deeper characterization of the identified phenotypes. This integrative approach may serve as a blueprint for uncovering molecular signatures of sepsis subgroups and holds promise for the development of future targeted therapies.

## Background

In recent decades, the major improvements in sepsis therapy were almost exclusively based on the quick application of an adequate infectiological treatment, and hemodynamic stabilization through fluid resuscitation and application of vasopressors (1). However, with high mortality rates (2), sepsis remains a major burden in global healthcare. Apart from infectious disease and symptomatic treatment, specific therapeutic approaches directly addressing the molecular alterations in sepsis, have consistently failed or shown contradictory results (3, 4). One possible explanation for these findings is seen in the fact, that clinical sepsis trials enrolled heterogeneous patient populations (5), leading to non-comparable sepsis manifestations. The identification of clinically relevant sepsis phenotypes might therefore be essential for the future development and predictive enrichment for specific sepsis therapies (6).

These days, a large amount of routine intensive care data is widely available, so that unsupervised machine learning (ML) methods appear to be well suited for identifying clinical sepsis phenotypes (7). However, clinical phenotyping relies on routine data which serve as surrogate parameters rather than capturing the true underlying biomolecular alterations in sepsis. As a result, the identified subtypes offer limited potential for informing novel, personalized therapeutic strategies. To date, there is only limited information on the biomolecular characteristics that are associated with clinical phenotypes, thus limiting the opportunities for development of therapeutic strategies (6). Moreover, since the course of sepsis is a highly dynamic process, temporal shifts in sepsis subtypes within individual patients may be a key factor in distinguishing critical from non-critical disease trajectories. Despite this potential, the temporal dimension of sepsis progression has so far rarely been studied and been largely neglected (8).

In this study, we aimed to (i) identify distinct sepsis phenotypes by unsupervised machine learning using routine clinical data, (ii) translate these subtypes to the molecular level to reveal biomolecular alterations relevant for personalized therapies by mass spectrometry-based plasma proteomics, (iii) analyze the trajectories of patients through these subtypes in accordance with their outcome, and (iv) develop and evaluate a supervised machine learning model that reliably detects these subtypes at the onset of sepsis treatment using a minimal set of universally accessible clinical variables, thereby laying the groundwork for a future clinical decision support system.

## Methods

### Patient cohort

This work is based on the multicenter, prospective, observational SepsisDataNet.NRW and CovidDataNet.NRW studies (German clinical trial registry, no. DRKS00018871). Patients were eligible for inclusion when treated for sepsis meeting sepsis 3 criteria (9) within the previous 48 hours at time of inclusion. In CovidDataNet.NRW patients were included when the inclusion criteria were complemented by an active SARS-CoV-2 infection. Patient-related ICU data from electronic health records was collected and blood samples were obtained regularly. Therapy was not affected by study inclusion and was based on latest sepsis guidelines (1). Parts of the cohort have already been analyzed in other contexts (10, 11).

### Cytokine measurements

The concentrations of 13 cytokines were quantified in serum samples using the LegendPlex Human Inflammation Panel 1 (Biolegend, San Diego) according to the manufacturer’s instructions. Briefly, the LegendPlex beads were incubated with serum samples, subsequently washed, incubated with detection antibodies, and washed again. Measurements were carried out using a flow cytometer (Canto II, BD Biosciences, CA) and concentration were interpolated from calibration curves. Values below the lower limit of quantification were considered to be 0 ng/mL, values higher than the upper limit of quantification were replaced with the respective upper limit of quantification.

### Clustering

Clinical variables on the day of sepsis diagnosis which had the following characteristics were selected for clustering: 1) routine assessment in the ICU, 2) less than 30% missing values 3) representation of the clinical severity of organ damage rather than an association with a treatment regime (Supplementary Table 1). For variables with multiple observations per day data was aggregated by the median, minimum or maximum, using the most clinical informative value (e.g. minimum value for white blood cell count and maximum value for creatinine). Missing values were imputed using data from the subsequent two days. If no corresponding values were available, the median of the respective variable was used for imputation. In addition to the clinical variables, the site of infection and the age of the patients were used for clustering. To adjust the weighting of different numerical values, the data was z-transformed before Principal Component Analysis (PCA) was carried out for dimension reduction. Different sepsis phenotypes were identified using the k-means algorithm on the PCA results of the first 11 components (>70% of variance). The number of clusters were identified using silhouette curves, finding three different clusters (Supplementary Figure 1). For the time series, the data from days 4, 7, and 9 were transformed using the standardization parameters (mean and standard deviation) of day 1 to ensure consistent transformation across the timepoints. The resulting principal components of day one were used to project the data from the subsequent days into the same reduced feature space. To assign the data from the following days to the clusters, the Euclidean distance to each cluster center was calculated, and each data point was assigned to the cluster with the nearest cluster center. Calculations were done using Python (v.3.11.5) and the packages pandas (v.2.1.1) and scikit-learn (v.1.3.1).

### Plasma Proteomics

A detailed description of plasma proteomics analyses can be found as supplementary material. Briefly, 1 µl plasma per sample was digested with trypsin according to the SP3 protocol as described before (12). 455 samples from 301 patients and two time points were analyzed by LC-MS/MS in setups using either an Orbitrap Fusion Lumos or an Exploris 240 mass spectrometer (Thermo Scientific, Bremen, Germany). Data were acquired in data-independent acquisition mode and analyzed using DIA-NN (v.1.8.1) with a spectral-library that was generated with FragPipe (v.18) searching the UniProt/SwissProt data base restricted to *Homo sapiens* (v.2022_05). Sample batches were processed individually and subsequently normalized for batch effects as described before (12) (Supplementary Figure 2). Differences in protein intensities between the clusters were tested for significance by ANOVA followed by Tukey’s post-hoc test. Proteins with a minimum of five observations per cluster were considered for testing. ANOVA p-values were corrected using the method of Benjamini-Hochberg. Proteins with an ANOVA p_FDR_ value and a post-hoc p-value ≤0.05 were considered significant. Functional annotation and enrichment analyses were carried out using the STRING web interface (string-db.org, v.12.0).

### Machine Learning

For machine learning, routine ICU data with less than 30% missing values were used (Supplementary Table 2). Data preprocessing, such as variable aggregation and imputation, was done using the same approach as in the clustering process. Before model development, highly correlated features were removed by calculating the Pearson correlation between features and excluding one of the two features with an absolute correlation greater than 0.7 (Supplementary Table 3). To select the most relevant features, a multiclass classifier based on a Random Forest model was employed. The data were split into training and test sets (70 % training, 30 % test), and a 100 times Monte Carlo Cross Validation (MCCV) (13) was performed to evaluate model metrics for different feature sets. The order of features for iterative training was determined based on an additional 100 times MCCV in the training dataset (70 % training, 30 % validation, Supplementary Figure 3). In this process, the median ranks of feature importance were computed using SHAP values (Shapley Additive Explanations). Missing values were imputed using the median in each MCCV iteration. Cluster C was bootstrapped to 60% of the size of Cluster A in every training set of each MCCV to prevent a bias towards the over-represented cluster. To determine the optimal number of features, the averaged recall was plotted against the number of selected features. The point at which the recall growth rate markedly declined was identified using the *KneeLocator* function of the keed package in Python. Tune Hyperparameters can be found in Supplementary Table 4. Calculations were done using Python (v.3.10.12) and the packages pandas (v.2.2.2), numpy (v.1.26.4), keed (v.0.8.5), shap (v0.46.0) and scikit-learn (v.1.5.1).

## Results

We identified three clusters representing different clinical sepsis phenotypes. These phenotypes showed a significant difference in the SOFA score (Supplementary Table 5), representing the *disease severity* of septic patients. Between the clusters, SOFA scores were gradually increasing from a median value of 7 (IQR 4-11) in cluster A (n = 202; 53 %), over 10 (6–13) in cluster B (n = 156; 41 %), to 15 (13–17) in cluster C (n = 26; 7 %). The baseline characteristics with a focus on different organ systems affected by sepsis are presented in Table 1.

**Table 1.**
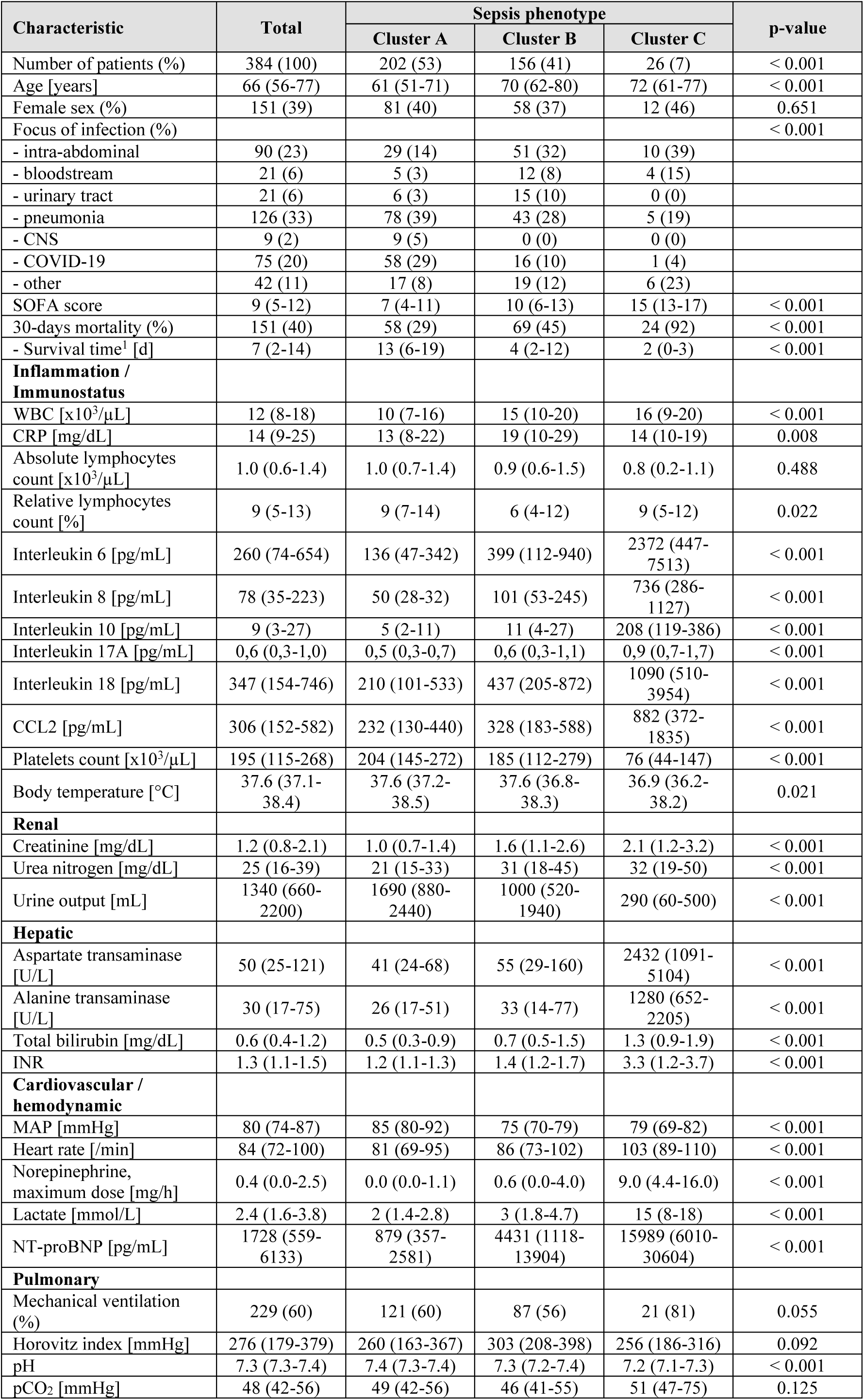
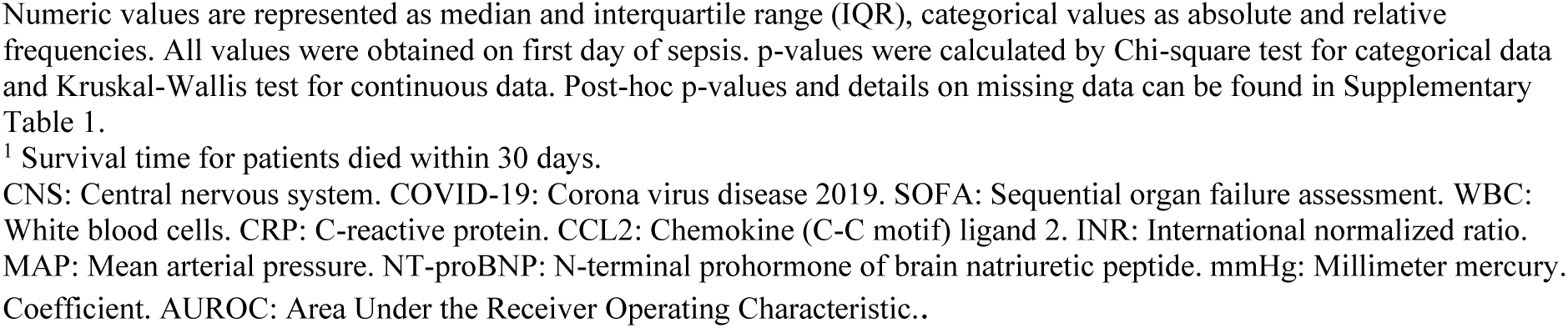
Baseline characteristics of the study cohort, differentiated by sepsis phenotype.

| Characteristic | Total | Sepsis phenotype |  |  | p-value |
| --- | --- | --- | --- | --- | --- |
|  |  | Cluster A | Cluster B | Cluster C |  |
| Number of patients (%) | 384 (100) | 202 (53) | 156 (41) | 26 (7) | < 0.001 |
| Age [years] | 66 (56-77) | 61 (51-71) | 70 (62-80) | 72 (61-77) | < 0.001 |
| Female sex (%) | 151 (39) | 81 (40) | 58 (37) | 12 (46) | 0.651 |
| Focus of infection (%) |  |  |  |  | < 0.001 |
| - intra-abdominal | 90 (23) | 29 (14) | 51 (32) | 10 (39) |  |
| - bloodstream | 21 (6) | 5 (3) | 12 (8) | 4 (15) |  |
| - urinary tract | 21 (6) | 6 (3) | 15 (10) | 0 (0) |  |
| - pneumonia | 126 (33) | 78 (39) | 43 (28) | 5 (19) |  |
| - CNS | 9 (2) | 9 (5) | 0 (0) | 0 (0) |  |
| - COVID-19 | 75 (20) | 58 (29) | 16 (10) | 1 (4) |  |
| - other | 42 (11) | 17 (8) | 19 (12) | 6 (23) |  |
| SOFA score | 9 (5-12) | 7 (4-11) | 10 (6-13) | 15 (13-17) | < 0.001 |
| 30-days mortality (%) | 151 (40) | 58 (29) | 69 (45) | 24 (92) | < 0.001 |
| - Survival time<sup>1</sup> [d] | 7 (2-14) | 13 (6-19) | 4 (2-12) | 2 (0-3) | < 0.001 |
| **Inflammation / Immunostatus** |  |  |  |  |  |
| WBC [$\times 10^3/\mu\text{L}$] | 12 (8-18) | 10 (7-16) | 15 (10-20) | 16 (9-20) | < 0.001 |
| CRP [mg/dL] | 14 (9-25) | 13 (8-22) | 19 (10-29) | 14 (10-19) | 0.008 |
| Absolute lymphocytes count [$\times 10^3/\mu\text{L}$] | 1.0 (0.6-1.4) | 1.0 (0.7-1.4) | 0.9 (0.6-1.5) | 0.8 (0.2-1.1) | 0.488 |
| Relative lymphocytes count [%] | 9 (5-13) | 9 (7-14) | 6 (4-12) | 9 (5-12) | 0.022 |
| Interleukin 6 [pg/mL] | 260 (74-654) | 136 (47-342) | 399 (112-940) | 2372 (447-7513) | < 0.001 |
| Interleukin 8 [pg/mL] | 78 (35-223) | 50 (28-32) | 101 (53-245) | 736 (286-1127) | < 0.001 |
| Interleukin 10 [pg/mL] | 9 (3-27) | 5 (2-11) | 11 (4-27) | 208 (119-386) | < 0.001 |
| Interleukin 17A [pg/mL] | 0,6 (0,3-1,0) | 0,5 (0,3-0,7) | 0,6 (0,3-1,1) | 0,9 (0,7-1,7) | < 0.001 |
| Interleukin 18 [pg/mL] | 347 (154-746) | 210 (101-533) | 437 (205-872) | 1090 (510-3954) | < 0.001 |
| CCL2 [pg/mL] | 306 (152-582) | 232 (130-440) | 328 (183-588) | 882 (372-1835) | < 0.001 |
| Platelets count [$\times 10^3/\mu\text{L}$] | 195 (115-268) | 204 (145-272) | 185 (112-279) | 76 (44-147) | < 0.001 |
| Body temperature [°C] | 37.6 (37.1-38.4) | 37.6 (37.2-38.5) | 37.6 (36.8-38.3) | 36.9 (36.2-38.2) | 0.021 |
| **Renal** |  |  |  |  |  |
| Creatinine [mg/dL] | 1.2 (0.8-2.1) | 1.0 (0.7-1.4) | 1.6 (1.1-2.6) | 2.1 (1.2-3.2) | < 0.001 |
| Urea nitrogen [mg/dL] | 25 (16-39) | 21 (15-33) | 31 (18-45) | 32 (19-50) | < 0.001 |
| Urine output [mL] | 1340 (660-2200) | 1690 (880-2440) | 1000 (520-1940) | 290 (60-500) | < 0.001 |
| **Hepatic** |  |  |  |  |  |
| Aspartate transaminase [U/L] | 50 (25-121) | 41 (24-68) | 55 (29-160) | 2432 (1091-5104) | < 0.001 |
| Alanine transaminase [U/L] | 30 (17-75) | 26 (17-51) | 33 (14-77) | 1280 (652-2205) | < 0.001 |
| Total bilirubin [mg/dL] | 0.6 (0.4-1.2) | 0.5 (0.3-0.9) | 0.7 (0.5-1.5) | 1.3 (0.9-1.9) | < 0.001 |
| INR | 1.3 (1.1-1.5) | 1.2 (1.1-1.3) | 1.4 (1.2-1.7) | 3.3 (1.2-3.7) | < 0.001 |
| **Cardiovascular / hemodynamic** |  |  |  |  |  |
| MAP [mmHg] | 80 (74-87) | 85 (80-92) | 75 (70-79) | 79 (69-82) | < 0.001 |
| Heart rate [/min] | 84 (72-100) | 81 (69-95) | 86 (73-102) | 103 (89-110) | < 0.001 |
| Norepinephrine, maximum dose [mg/h] | 0.4 (0.0-2.5) | 0.0 (0.0-1.1) | 0.6 (0.0-4.0) | 9.0 (4.4-16.0) | < 0.001 |
| Lactate [mmol/L] | 2.4 (1.6-3.8) | 2 (1.4-2.8) | 3 (1.8-4.7) | 15 (8-18) | < 0.001 |
| NT-proBNP [pg/mL] | 1728 (559-6133) | 879 (357-2581) | 4431 (1118-13904) | 15989 (6010-30604) | < 0.001 |
| **Pulmonary** |  |  |  |  |  |
| Mechanical ventilation (%) | 229 (60) | 121 (60) | 87 (56) | 21 (81) | 0.055 |
| Horovitz index [mmHg] | 276 (179-379) | 260 (163-367) | 303 (208-398) | 256 (186-316) | 0.092 |
| pH | 7.3 (7.3-7.4) | 7.4 (7.3-7.4) | 7.3 (7.2-7.4) | 7.2 (7.1-7.3) | < 0.001 |
| pCO<sub>2</sub> [mmHg] | 48 (42-56) | 49 (42-56) | 46 (41-55) | 51 (47-75) | 0.125 |

<sup>1</sup> Survival time for patients died within 30 days.
CNS: Central nervous system. COVID-19: Corona virus disease 2019. SOFA: Sequential organ failure assessment. WBC: White blood cells. CRP: C-reactive protein. CCL2: Chemokine (C-C motif) ligand 2. INR: International normalized ratio. MAP: Mean arterial pressure. NT-proBNP: N-terminal prohormone of brain natriuretic peptide. mmHg: Millimeter mercury. Coefficient. AUROC: Area Under the Receiver Operating Characteristic..

### Clinical representations of clusters

Least affected patients were found in cluster A, characterized by a 30-day mortality rate of 29 % (n = 58/202) with a median survival time of 13 days (IQR 6-19). The *low* mortality in this group was clinically underpinned by just a moderate organ failure and moderate vasopressor support. Patients who were allocated to this cluster mostly had either a sepsis due to pneumonia or COVID-19. Moreover, cluster A included the youngest patients with a median age of 61 years (IQR 51-71).

Cluster B represented a sepsis phenotype between most and least affected patients. For patients allocated to cluster B on day 1 the median 30-day mortality rate was 45% (n = 69/156), with a median survival time of 4 days (IQR 2-12). The most common focus of infection in this cohort was located intra-abdominal. The main difference was the presence of just a moderate AKI. In comparison to cluster A, a higher rate of septic shock was evident. The most affected patients were allocated to cluster C, which also showed up in a markedly high 30-day mortality rate of 92 % (n = 24/26) with a median survival time of just 2 days (IQR 0-3). The cohort was suffering from multi-organ failure with a leading acute liver failure, lactate acidosis and accompanying high-grade acute kidney injury (AKI) with circulatory shock. The median age was 70 years (IQR 61-77).

The longitudinal analysis revealed substantial dynamics between clusters, with Cluster B exhibiting the highest variability (Figure 1). Notably, 39.7 % of patients initially assigned to Cluster B transitioned to a different state within three days—either to the more favorable Cluster A (35.3 %), the less favorable Cluster C (4.5 %), or due to a fatal outcome (14.7 %) (Supplementary Table 6). Cluster C showed a strong association with mortality with 97 % of patients (all but one) migrating through it eventually dying within the 30-day period.

**Fig. 1.**
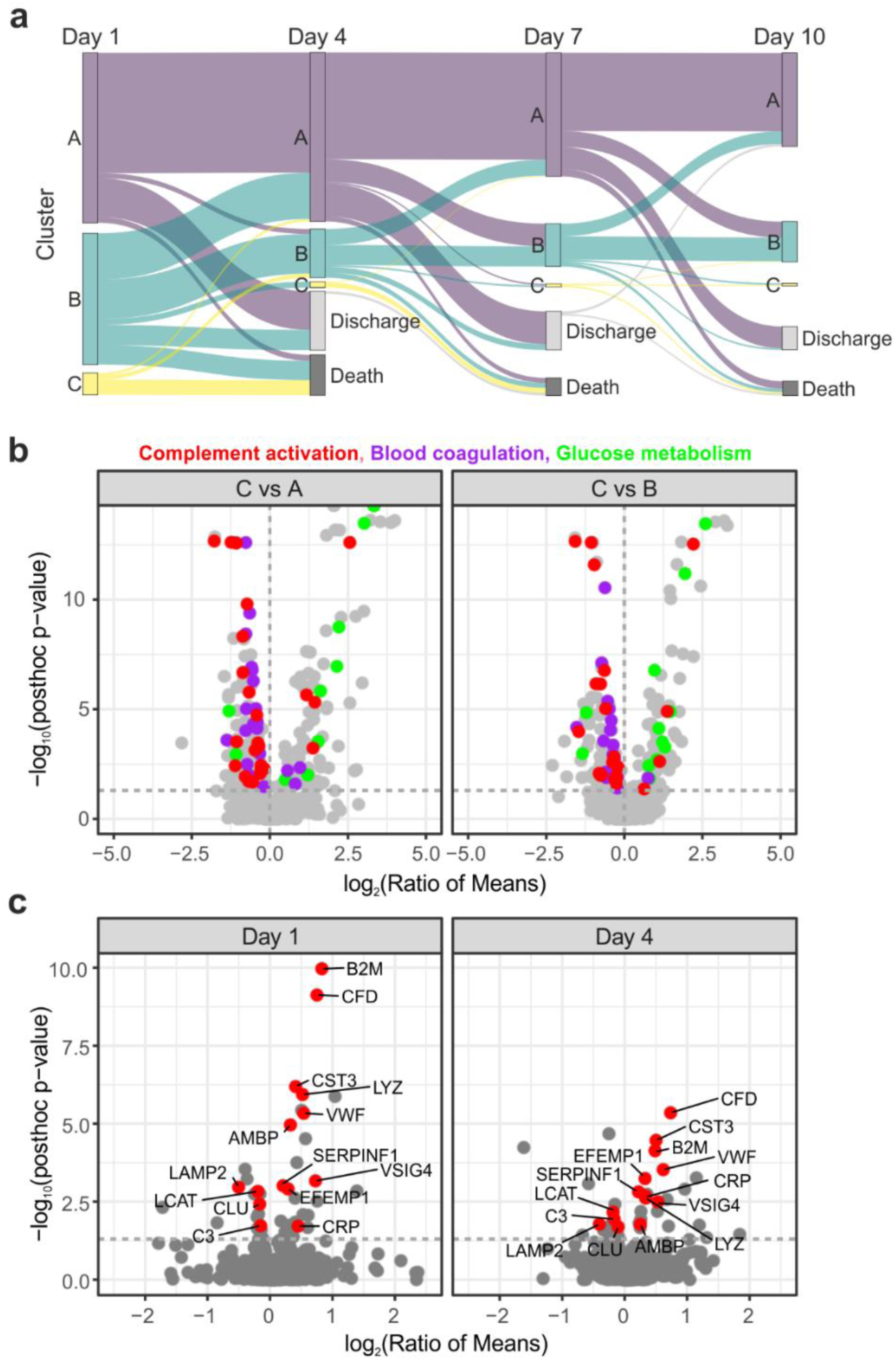
Time course analysis of clinical sepsis subtypes. **(a)** Sankey diagram illustrating the assignment of patients to clinical subtypes on days 1, 4, 7 and 10. Patients migrate between clusters or drop out of the analysis due to ICU discharge or fatal events. **(b)** Volcano plots illustrating the analysis of phenotypes A and B compared to C using ANOVA (Benjamini-Hochberg-corrected) followed by Tukey’s post-hoc test. Significantly differentially abundant proteins (p_FDR_ value ≤ 0.05, post-hoc p value ≤ 0.05) are annotated with the GO biological processes as indicated on the top. **(c)** Volcano plots showing the comparison of plasma proteome data between clusters A and B on day 1 and 4, respectively. Proteins that were found significant at both time points highlighted and labeled with gene names.

### Immunostatus

The immunological status of the sepsis phenotypes, showed the lowest relative lymphocyte count in cluster B with 6 % (4–12), compared to clusters A and C with 9 % each (IQR 7-14 and 5-12, respectively). Interleukins 6, 8, 10, 17A, and 18 representing the inflammatory status of patients had the highest level in cluster C and the lowest in cluster A. This was also observed for chemokine (C-C motif) ligand 2 (CCL2) with levels of 232 pg/mL (130–440), 328 (183–588), and 882 (372–1835) for clusters A, B, and C, respectively.

### Plasma proteome characteristics of cluster C

We observed massive changes in the plasma proteome of cluster C in comparison to the other two clusters. Overall, we quantified 609 plasma proteins (Supplementary Figure 4) of which at day 1 181 and 171 proteins were found significantly altered between clusters A and C and clusters B and C, respectively (Fig. 1b, Supplementary Table 7). The overlap between these proteins was 84% indicating specific proteome characteristics of cluster C. The differentially abundant proteins were associated with complement activation, blood coagulation and acute phase immune response (Figure 1b, Figure 2a). Strikingly, the related proteins were mainly lower abundant pointing towards an excessive consumption of complement and coagulation factors as well as acute phase proteins in the most affected patients (Figure 2b). Another major part of these protein alterations was associated with liver damage. We observed a striking enrichment of proteins related to glucose metabolism, probably released by liver cell death. The intensities of several proteins including fructose-bisphosphate aldolase B (ALDOB) and transaldolase (TALDO1) correlated with AST/ALT laboratory measurements (Figure 2c). The association of these proteins with canonical glycolysis pointed towards a shift towards anaerobic metabolism in the liver. In addition, a limited synthesis capacity of the injured liver probably also contributed to the decreased abundance of complement, coagulation and acute phase proteins. At day 4, much less proteins were found to be differentially abundant in comparison to cluster C, which was a result of its small size with only five patients in the proteome analysis (57 proteins were altered between clusters A and C and 51 proteins between clusters B and C, Supplementary Table 7). Still, proteins related to blood coagulation and complement activation were prominently regulated and markers of liver injury were characteristic for cluster C. In addition, Myoglobin (MB) was substantially elevated in cluster C, indicating damage of muscle tissue (14), that was less prominent at day 1.

**Fig. 2.**
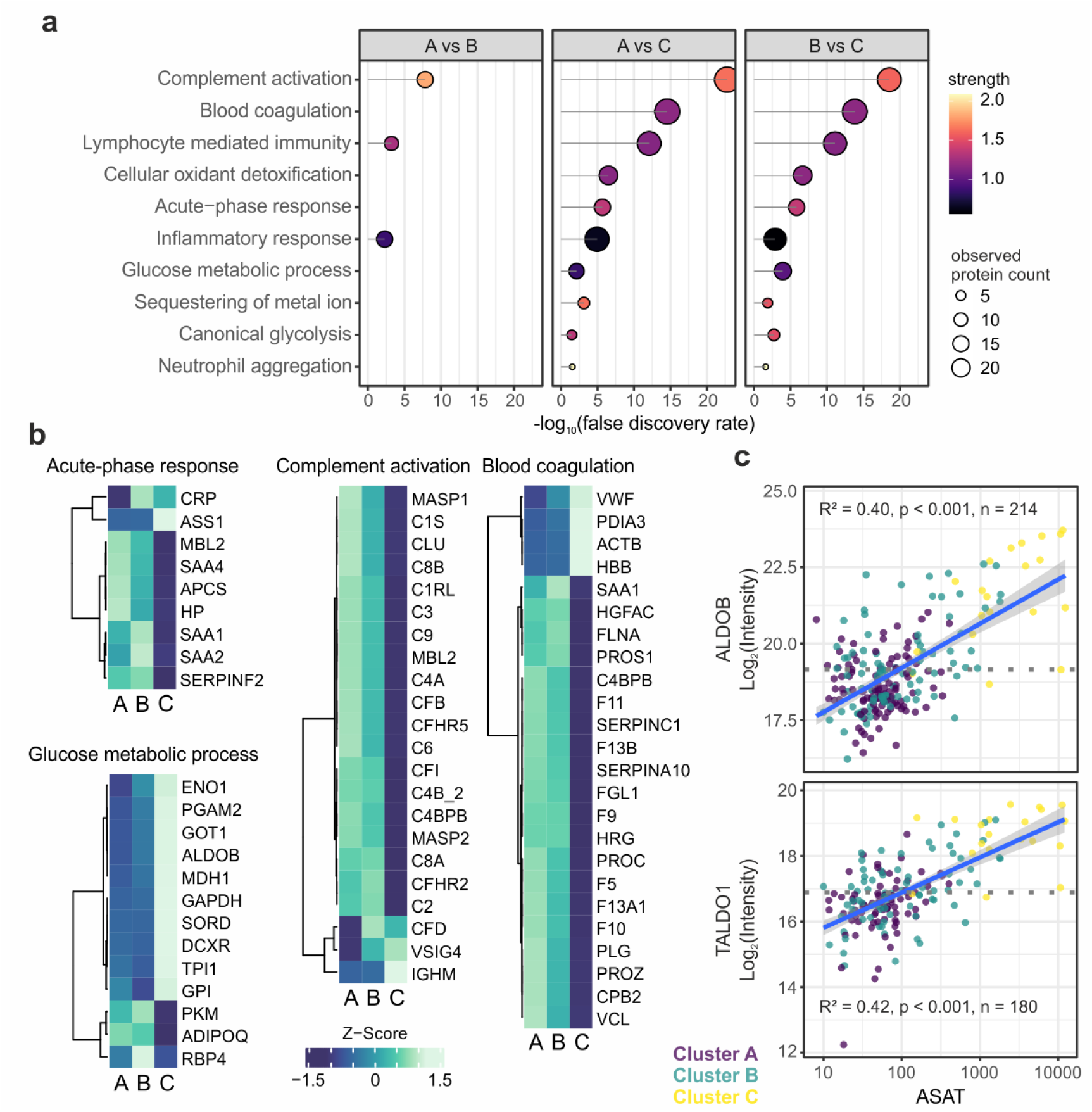
Proteomics characterization of clinical sepsis phenotypes at day 1. **(a)** Functional enrichment analysis based on GO biological processes. Analyses were performed separately for significantly differentially abundant proteins from the respective comparisons. Ten selected ontology terms are shown. **(b)** Heatmaps showing the mean intensities (z-scored) in the three phenotypes for all proteins, (Pearson correlation, complete linkage). **(c)** Scatter plots illustrating the linear regression analysis of protein intensities with aspartate aminotransaminase (ASAT) laboratory measurements. Blue line representing the linear fit with its confidence interval.

### Plasma proteome characteristics of clusters A and B

Clusters A and B showed less prominent differences with 40 differentially abundant proteins at day 1 and 29 proteins at day 4 (Supplementary Table 7, Figure 1c). Several regulated proteins reflected the difference in disease severity between the clusters. The V-set and immunoglobulin domain-containing protein 4 (VSIG4), which was described to be secreted by activated peritoneal macrophages (15), for instance showed a significant correlation with the SOFA score and was consequently higher abundant in cluster B. Complement factor 3 (C3), on the other hand, was inversely correlated with the SOFA score (Fig. 3a). Other proteins reflected AKI as also indicated by increased serum creatinine levels in cluster B. The low-molecular-weight proteins complement factor D (CFD) and beta-2 microglobulin (B2M), which normally freely pass the glomerular filtration, were significantly increased in cluster B (Fig. 3b) and correlated with the established marker of renal function, Cystatin C (CST3), as well as with each other (Fig. 3c). Importantly, these proteins were found differentially abundant at both time points. Cluster A showed the mildest sepsis phenotype which was associated with the highest levels of complement and coagulation factors (Fig.2b). This indicated the consumption of the corresponding proteins, which was already evident in cluster B and became excessive in cluster C.

**Fig. 3.**
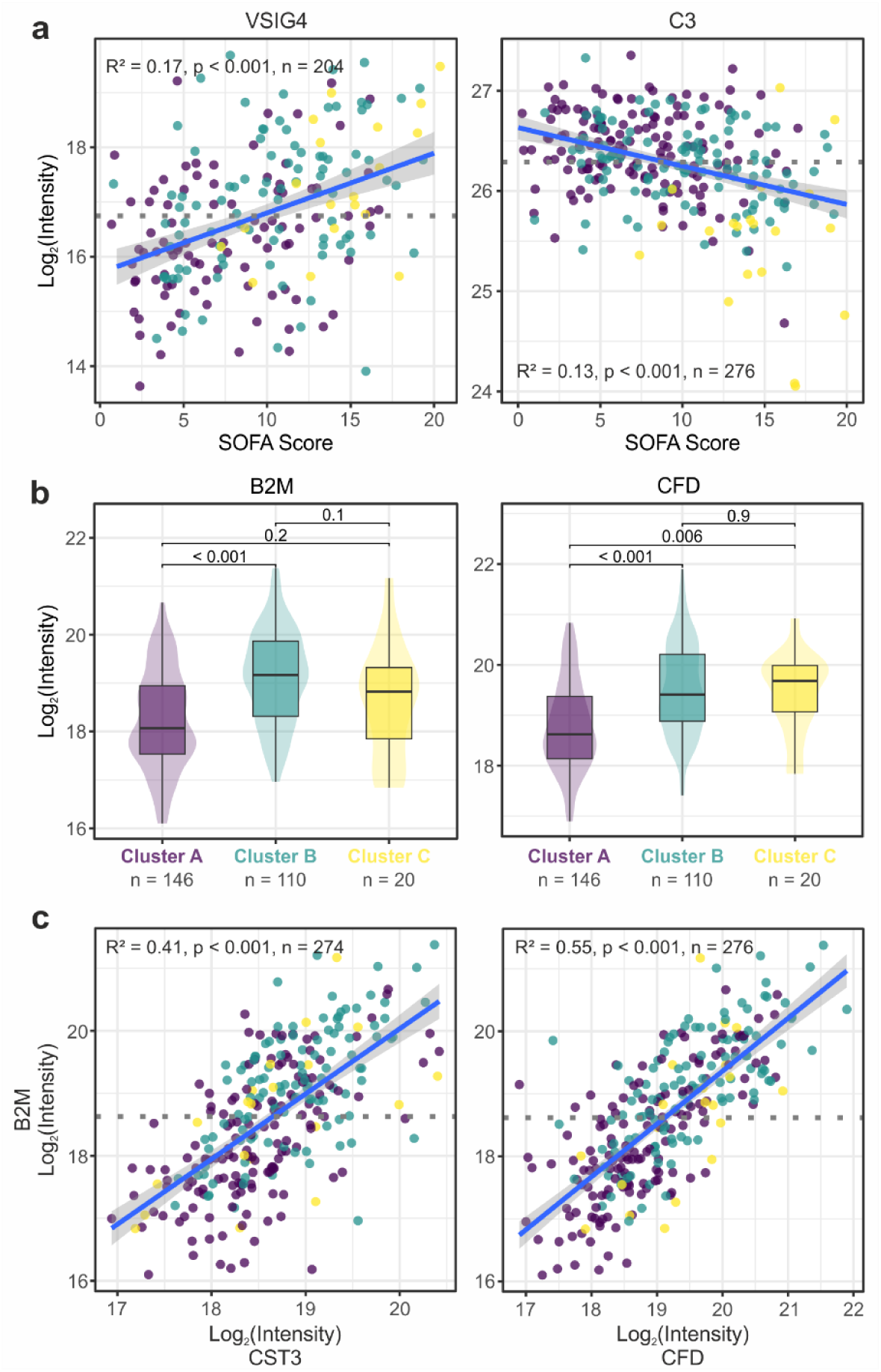
Comparison of clusters A and B at day 1. **(a)** Linear regression of V-set and immunoglobulin domain-containing protein 4 (VSIG4) and Complement factor 3 (C3), both significantly differentially abundant between cluster A and B, with the SOFA score. Colors of data points indicate the cluster the respective patients belong to. Blue line representing the linear fit with its confidence interval. **(b)** Boxplots illustrating the protein intensities of beta-2 microglobulin (B2M) and Complement factor D (CFD) in the three clusters. Boxes represent 25th and 75th percentiles, whiskers extend to the most extreme data points, median shown as a horizontal line, p-values from Tukey’s post-hoc test. **(c)** Linear regression analysis of B2M with Cystatin-C (CST3) and CFD.

### Patient classification by supervised machine learning

To enable a future stratification of patients and to identify the most relevant features for discrimination between the three clinical phenotypes, we trained a random forest ML classifier and performed feature importance analysis. By iteratively adding features to the which only marginal improvement in recall was achieved (Figure 4a). Over 100 iterations of Monte Carlo cross validation, the most frequently selected features in combinations with seven features were alanine transaminase (ALT), aspartate transaminase (AST), base excess (BE), international normalized ratio of thrombin time (INR), diastolic arterial blood pressure, systolic arterial blood pressure (BPdia, BPsys) and activated partial thromboplastin time (aPTT), all parameters which are widely available in clinical routine (Figure 4b, Supplementary Table 8). Across all evaluated metrics, a discrepancy greater than 0.1 was observed between the training and test sets, indicating overfitting, likely due to an insufficient number of data points (Supplementary Table 9). Despite overfitting, an acceptable model performance can still be achieved for unseen data across all three classes with an AUROC of 0.95 ± 0.016 as well as a precision of 0.813 ± 0.050 and a recall of 0.839 ± 0.043 (Figures 4c and d, Supplementary Table 9). These values should be interpreted with care, but they may serve as a proof-of-concept for future applications, demonstrating that the clusters can be predicted using only a few routinely collected features. Shapley feature importance analysis allows us to determine how individual features influence model predictions. In Cluster C, AST and ALT levels were elevated, while Clusters A and B differed in BE values, with higher BE levels favouring predictions for Cluster A (Supplementary Figure 5). Furthermore, this approach provides insights into how specific features contribute to the prediction of a cluster for a given patient (Supplementary Figure 6).

**Fig. 4.**
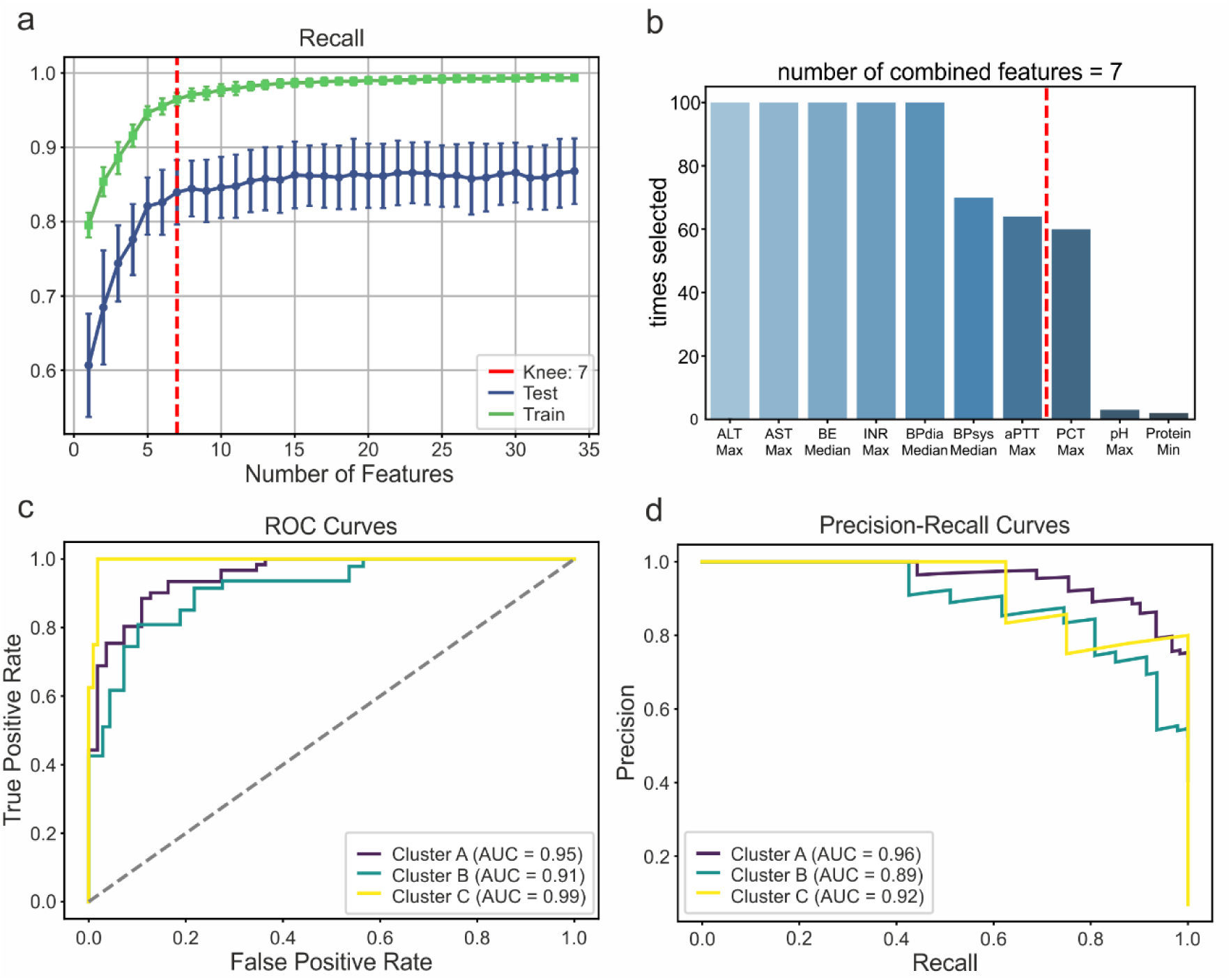
Model Performance of the Random Forest ML Classifier **(a)** Mean recall of the train and test data as a function of the number of features. The red dotted line indicates the computed knee point, where the slope decreases significantly at 7 features. **(b)** Bar chart showing the frequency at which a feature was included in a combination of seven features during MCCV. **(c)** Receiver Operating Characteristic (ROC) curves for all clusters, each compared to the rest, shown exemplarily from a single iteration during MCCV. (d) Precision-Recall curves for all clusters, each compared to the rest, shown exemplarily from a single iteration.

## Discussion

In the present study, we identified three clinical sepsis phenotypes and their temporal development in a prospective multicenter cohort using machine learning. The phenotypes represented different states of sepsis severity and differed in their clinical characteristics, the affected organ systems, and their immune status. Longitudinal analysis revealed dynamic characteristics of cluster B and a strong association with mortality of cluster C. We applied plasma proteomics to characterize the associated molecular alterations and found that the phenotypes translated to the plasma proteome. We could show how organ damage affected the composition of the plasma proteome and that components of the innate immune system depleted with increasing sepsis severity. Finally, we demonstrate that by using machine learning we are able to classify new patients by using just seven widely available clinical parameters while also interpreting the decision making.

Despite intensive research over the last decades, little progress has been made in sepsis therapy. High mortality and morbidity rates continue to bear witness to this (2). Sepsis is based on an infection, but organ damage is an expression of pronounced dysregulation of the immune system and other metabolic processes (16). Therefore, the most promising approach for future sepsis therapy is seen in the individualized and targeted treatment of metabolic and immunological alterations (17). To advance precision medicine in sepsis, it is essential not only to identify clinical phenotypes but also to understand the biological mechanisms that underlie them. Previous work, such as by Seymour et al., has successfully stratified sepsis patients into clinical subtypes using machine learning on routine data (7). However, these phenotypes were primarily based on surrogate markers (e.g., routine laboratory values) and lacked molecular characterization. In contrast, our study bridges this critical gap by integrating high-throughput plasma proteomics, providing biological characterization to phenotypes defined by clinical routine data (18). While direct proteomic phenotyping is currently not feasible at the bedside, we demonstrate that surrogate clinical phenotypes can offer meaningful biological insights if they are thoroughly characterized at the molecular level. This enables a biologically informed interpretation of clinical phenotypes and the early prediction of molecularly distinct disease courses using a minimal set of widely available clinical variables (19–22), which lays the foundation for hypothesis-driven stratification in interventional trials.

To give a concrete example: In the plasma proteome of patients most severely affected by sepsis (cluster C), we found a deficiency in complement and coagulation factors to a previously unknown extent (23, 24). While the coagulation system is well monitored by clinical measures, the complement system is not directly represented by routine data and its importance might be underestimated. This was underlined by the observation that complement factors were significantly reduced already in cluster B. Complement factor 3, a central hub of the complement cascade, showed a linear correlation with the SOFA score indicating a direct relation to sepsis severity rather than a secondary effect caused by impaired synthesis resulting from liver damage. Acute kidney injury, on the other hand, lead to the accumulation and elevated levels of low-molecular-weight plasma proteins, such as Complement factor D (CFD). CFD is the rate-limiting enzyme of the alternative pathway (AP) which is also responsible for signal amplification via the amplification loop (25). Thus, the accumulation of CFD may further escalate the excessive activation of the complement system, which is also known to cause tissue damage and thrombo-inflammation (26). Although the relevance of the complement is well established for COVID-19 (27) it is still largely overseen in polymicrobial sepsis and might represent a future target for therapeutic measures. Targeting CFD in cluster B would be an obvious strategy and inhibition of the AP amplification loop might benefit from limiting the destructive power of the complement cascade while preserving its basal activity and important functions such as leukocyte recruitment and activation (25).

Our findings suggest that identifying patients with specific biological profiles at the onset of sepsis may help to revive or refine targeted therapies. In this sense, longitudinal analysis of phenotypes is of great importance, as the time at which patients are assigned to phenotypes is influenced by many factors. Our data suggest that phenotypes in sepsis are stable over a significant period of time and show that this is also the case at the plasma proteome level. This observation is important if we hypothesize that clinical decision making should be supported by clinical phenotypes. On the other hand, we observed changes in phenotypes in a subset of patients that were associated with mortality when they migrated to cluster C, demonstrating that phenotypes could also help to monitor patients over the course of therapy. Importantly, we show that within these dynamics the molecular profile of the plasma proteome was consistent between phenotypes and the examined time points. Thus, our study not only provides a classification framework but also a biological rationale for future precision-guided clinical interventions (18). Moreover, our machine learning models enable the simple allocation of patients to clinical phenotypes, offering a pragmatic approach to anticipate molecular profiles and guide phenotype-informed interventions.

### Limitations

The strength of our study result from the prospective nature of the patient inclusion, so that unlike the retrospective use of secondary data, our recruited patients were certain to have sepsis. To the best of our knowledge, this is the largest cohort of sepsis patients that have been characterized using machine learning in combination with plasma proteomics. The limiting factor is that our cohort is small compared to studies that have performed clustering on sepsis patients, so that the transferability of the results cannot necessarily be generalized. It is therefore imperative that our results are validated in further cohorts. Further limitations are that due to the unique composition of the plasma proteome and technical limitations of LC-MS/MS, it is currently extremely challenging to measure low abundant proteins, such as certain cytokines. Thus, our observations are limited to the top 600 plasma proteins that were covered by the present analysis.

### Conclusions

In this study, we identified distinct clinical sepsis phenotypes that not only differed in disease severity, progression and outcomes, but also showed characteristic patterns in the plasma proteome, reflecting underlying metabolic and immunological alterations. These findings emphasize the potential of combined clinical and molecular phenotyping to uncover biologically meaningful subgroups in sepsis. Such stratification could be critical to guide the development and re-evaluation of targeted therapies. Our results highlight the importance of incorporating temporal dynamics and molecular data into sepsis research to improve patient stratification. While further validation in larger cohorts is required, our approach offers a blueprint for future studies aiming to translate biological insights into clinical decision-making. We advocate for a collaborative, multi-omics, and longitudinal research strategy to ultimately enable precision medicine in sepsis and design more effective, phenotype-guided interventional trials.

## Data Availability

The mass spectrometry proteomics data have been deposited to the ProteomeXchange Consortium via the PRIDE partner repository with the dataset identifier PXD058562.

## List of abbreviations

AKI: Acute kidney injury
ALT: Alanine transaminase
AST: Aspartate transaminase
AP: Alternative pathway of complement activation
aPTT: Activated partial thromboplastin time
COVID-19: Coronavirus disease 2019
FDR: False discovery rate
ICU: Intensive care unit
INR: International normalized ratio
LC-MS/MS: Liquid chromatography-coupled tandem mass spectrometry
MCCV: Monte Carlo cross validation
ML: Machine learning
PCA: Principal Component Analysis
SARS-CoV-2: Severe acute respiratory syndrome coronavirus type 2
SOFA: Sepsis-related organ failure assessment score

## Declarations

### Ethics approval and consent to participate

The SepsisDataNet.NRW and CovidDataNet.NRW studies were approved by the Ethics Committee of the Medical Faculty of Ruhr-University Bochum (Registration No. 5047–14 and 19-6606_6-BR, respectively) or the responsible ethics committee of each respective study center and conducted in accordance with the revised Declaration of Helsinki. Written informed consent was obtained from the patients or a legal representative.

### Consent for publication

Not applicable

### Availability of data and materials

Reviewer access details

Log in to the PRIDE website using the following details:

Project accession: PXD058562

Token: UGnQcBpmig7a

Alternatively, reviewer can access the dataset by logging in to the PRIDE website using the following account details:

Username

Password: 90Lvu7du6d5k

### Author contributions

Conceptualization: TB, MS, BS, HN, Writing the original draft: TB, KK, MW, NH, LP, Revision of original draft: MB, KS, TR, BK, KR, DZ, DH, AZ, TG, ME, BS, MA, Data generation & patient recruitment: LP, MB, KF, TR, MU, HH, AW, BK, KR, TG, DZ, UL, DH, SFE, AZ, Data analysis: TB, KK, MW, KS, HN, Supervision: MA, BS

## Acknowledgments

We thank the SepsisDataNet.NRW research group (alphabetical order): Michael Adamzik^1^, Moritz Anft^8^, Thorsten Annecke^5^, Nina Babel^8^, Maha Bazzi^1^, Lars Bergmann^1^, Christian Bode^4^, Thilo Bracht^9^, Alexander von Busch^1^, Jerome M Defosse^5^, Stefan F. Ehrentraut^4^, Martin Eisennacher^2^, Björn Ellger^7^, Christian Ertmer^3^, Ulrich H. Frey^9^, Katrin Fuchs^9^, Helge Haberl^1^, Dietrich Henzler^6^, Daniel Kleefisch^2^, Thomas Köhler^6^, Björn Koos^1^, Ulrich Limper^5^, Katrin Marcus^9^, Hartmuth Nowak^1^, Daniel Oswald^7^, Christian Putensen^4^, Tim Rahmel^1^, Katharina Rump^1^, Jens-Christian Schewe^4^, Elke Schwier^6^, Barbara Sitek^2^, Matthias Unterberg^1^, Frank Wappler^5^, Katrin Willemsen^1^, Alexander Wolf^1^, Alexander Zarbock^3^, Birgit Zuelch^1^

^1^Klinik für Anästhesiologie, Intensivmedizin und Schmerztherapie, Universitätsklinikum Knappschaftskrankenhaus Bochum, Bochum, Germany

^2^ Ruhr Universität Bochum, Medizinische Fakultät, Medizinisches Proteom-Center, Bochum, Germany.

^3^ Klinik für Anästhesiologie, operative Intensivmedizin und Schmerztherapie, Universitätsklinikum Münster, Münster, Germany

^4^ Klinik für Anästhesiologie und operative Intensivmedizin, Universitätsklinikum Bonn, Bonn, Germany

^5^ Klinik für Anaesthesiologie und operative Intensivmedizin, Universität Witten/Herdecke, Krankenhaus Köln-Merheim, Köln, Germany

^6^ Klinik für Anaesthesiologie und operative Intensiv-, Rettungsmedizin und Schmerztherapie, Klinikum Herford, Herford, Germany

^7^ Klinik für Anästhesiologie, Intensivmedizin und Schmerztherapie, Klinikum Westfalen, Dortmund, Germany

^8^ Centrum für Translationale Medizin, Medizinische Klinik I, Marien Hospital Herne, Universitätsklinikum der Ruhr-Universität Bochum, Herne, Germany.

^9^ Klinik für Anästhesiologie, operative Intensivmedizin, Schmerz-und Palliativmedizin, Marien Hospital Herne, Universitätsklinikum der Ruhr-Universität Bochum, Bochum, Germany

