## Supplementary Material for "Machine learning identifies clinical sepsis phenotypes that translate to the plasma proteome: a prospective cohort study"

**Table of contents**


| | |
|---|---|
| **1** | Supplementary Methods |
| **2** | Supplementary Table 1 Clinical features that were used for k-means clustering. |
| **3** | Supplementary Table 2 Clinical features that were used for machine learning. |
| **4** | Supplementary Table 3 Correlating feature pairs in the clinical dataset as evaluated by the Pearson Correlation Coefficient. |
| **5** | Supplementary Table 4: Hyperparameters for the random forest models for the prediction with clinical and proteomics data. |
| **6** | Supplementary Table 6: Individual patients and their assignment to cluster A, B and C across the studied time points. |
| **7** | Supplementary Table 7 Significantly differentially abundant proteins as determined by plasma proteomics. |
| **8** | Supplementary Table 8: Machine learning model performance metrics. |
| **9** | Supplementary Table 9 Feature importance ranks for random forest classifier. |
| **10** | Supplementary Figure 1: Silhouette curves that were used for determination of the optimal number of clusters. |
| **11** | Supplementary Figure 2: Quality control of proteomics batch normalization. |
| **12** | Supplementary Figure 3: Framework for Machine Learning and Feature Selection. |
| **13** | Supplementary Figure 4: Illustration of the quantified plasma proteome. |
| **14** | Supplementary Figure 5: Exemplary SHAP summary plots. |
| **15** | Supplementary Figure 6: Exemplary SHAP waterfall plots. |

**Supplementary Methods**

*Proteomics Sample Preparation*

All chemicals were purchased from Sigma-Aldrich or Thermo Fisher Scientific unless otherwise indicated. 1 µl plasma per sample was diluted 1:24 (v/v) in 100 mTris(hydroxymethyl)aminomethane pH 8.5, 10 mM Tris(2-carboxyethyl)phosphine, 40 mM 2-Chloroacetamide and 1% sodium deoxycholate and incubated for 10 min at 95 °C. Subsequently, 28 µL ammonium bicarbonate (Ambic, 50 mM) as well as 400 µg paramagnetic Sera-Mag Carboxyl-Magnet-Beads (Beads, Cytiva, GE Healthcare, Chicago, IL) were added. Acetonitrile (ACN) was added to a final concentration of 70 %, following 18 min incubation with periodical vortexing. Then, the beads were washed twice with 70 % ethanol and once with ACN. Remaining ACN was allowed to evaporate following overnight digestion of proteins using 1.5 µg trypsin (SERVA Electrophoresis, Heidelberg, Germany) in 50 mM Ambic at 37 °C. The resulting peptides were transferred to a new reaction tube and the digestion was stopped by adding 10 % trifluoroacetic acid (TFA) to a final concentration of 0.1 % (v/v).

*LC-MS/MS Analysis*

Samples from 276 patients were analyzed distributed over eight batches using three different LC-MS setups. Batch S3 was measured using an Ultimate 3000 RSLCnano HPLC coupled an Orbitrap Fusion Lumos mass spectrometer (both Thermo Scientific, Bremen, Germany). The peptides were preconcentrated for 7 minutes at a flow rate of 30 µl/min on a trap column (Acclaim PepMap 100, 75 µm × 2 cm, C18, 5 µm, 100 Å) using 0.1 % TFA. The separation was subsequently performed on an analytical column (Acclaim PepMap RSLC, 75 µm × 50 cm, nano Viper, C18, 5 µm, 100 Å) with a gradient of 5-30 % solvent B over 38 min at a flow rate of 400 nl/min and a column temperature of 60 °C (solvent A: 0.1 % formic acid (FA); solvent B: 0.1 % FA, 84 % ACN). The Fusion Lumos was operated in DIA mode with 21 windows between 350 and 1400 m/z. MS1 resolution was set to 60k with a RF lens setting of 30 %, a maximum injection time (MIT) of 80 ms and an AGC (automatic gain control) target of $4e^5$. For MS2 scans the ions were fragmented using HCD with a normalized collision energy (nCE) of 28 %. The resolution was set to 30k in a m/z range from 372 to 1275. MIT was set to 54 ms and AGC to 375000. Batches C3 and S9 were analyzed using a Vanquish Neo UHPLC coupled to an Orbitrap Exploris 240. Here, the separation was achieved with a flow rate of 0.5 µL per min, a column temperature of 60 °C and a gradient from 1 % B to 30

% B within the first 50 min, to 24 % B in another 2 min and finally to 45 % B in 3 min. Then, a washing step with alternating concentrations of 99 % and 1% B was programmed for 14 min. Contrary to the setups with Ultimate 3000 RSLCnano HPLCs a concentration of 80 % ACN in 0.1 % FA was used as solvent B. The Exploris 240 was operated in DIA mode with 23 windows between 380 and 1380 m/z. MS1 resolution was set to 120k, the RF Lens to 85 %, AGC to $1e^6$ and MIT to auto. For MS2 the HCD nCE was set to 30 %, the scan range to 145-1450 m/z, the resolution to 30k, the AGC target to $1e^6$ and the MIT to 80 ms. All other batches were analyzed using an Ultimate 3000 RSLCnano HPLC coupled to an Orbitrap Exploris 240. Here, the MS settings were as previously stated. For the separation of the analytes either a 15 cm self-packed analytical column (C18, ReproSil-Pur (Dr. Maisch HPLC GmbH, Ammerbuch, Germany), 75 µm x 15 cm, 1.9 µm particle size, 120 Å pore size) with a flow rate of 400 nL/min or a DNV PepMap 1500 bar C18 analytical column (75 mm x 150 mm, 2 µm particle size, 100 Å pore size, Thermo Fisher Scientific) with a flow rate of 500 nL/min were used. The separation was carried out using a 40 min gradient of 1-35 % solvent B. Here, 1 % solvent B was maintained for the first 7 minutes, followed by two increasing steps to 25 % solvent B for 28 minutes and afterwards to 35 % B in 5 minutes. The columns were heated to 60°C.

*Proteomics Data Analysis*

A spectral library was generated using DDA data which was generated using a Fusion Lumos or a QExactive HF instrument as described before (Unterberg et al., Crit Care. 2023 Oct 31;27(1):417). The respective raw data was processed using FragPipe (v.18) operated with default settings and searching the UniProt/SwissProt data base restricted to *Homo sapiens* (v.2022_05). The DIA data was processed with DIA-NN (ver.1.8.1) using the spectral library and data base mentioned before. Deep learning was activated and default parameters were used, expect for heuristic protein inference, which was disabled and the network classifier which was set to double pass mode. All batches were processed individually and normalized by the software (RT-dependent). Subsequently, the resulting data sets were imported into R and merged by the first accession of the protein groups. The protein intensities of each batch were first log2-transformed and normalized separately using the loess method (Välikangas et al., Brief Bioinform. 2018 Jan 1;19(1):1-11.). Then a linear model was calculated for each protein that estimated the batch effect for each batch. This batch effect was then subtracted from the protein intensities of the corresponding batch to reduce the differences between batches. The quality of the batch normalization was assessed using boxplots, principal component analysis (PCA) plots, heatmaps and MA plots. Differences in protein intensities between clusters were tested for statistical significance by ANOVA followed by Tukey's post-hoc tests. Proteins with a minimum of five observations per cluster were considered for testing. ANOVA p-values were corrected using the method of Benjamini-Hochberg. Ratios of mean intensities were calculated based on delogarithmized intensities. Proteins with an ANOVA pFDR value and a post-hoc p-value $\leq 0.05$ were considered significant. Functional annotation and enrichment analyses were carried out using the STRING web interface (string-db.org, v.12.0).

### Supplementary Tables

**Supplementary Table 1:** Clinical features that were used for k-means clustering.

| # | Abbreviation | Description | Location parameter |
|---|---|---|---|
| 1 | BE_MEDIAN | base excess (arterial blood gas) | median |
| 2 | BR_total_MAX | total bilirubin level (serum) | maximum |
| 3 | CL_MIN | chlorine level (serum of point-of-care testing) | minimum |
| 4 | GGT_MAX | gamma-glutamyltransferase level (serum) | maximum |
| 5 | AST_MAX | aspartate transaminase level (serum) | maximum |
| 6 | ALT_MAX | alanine transaminase level (serum) | maximum |
| 7 | HR_MEDIAN | heart rate | median |
| 8 | HGB_MIN | hemoglobin level (blood) | minimum |
| 9 | BP_mean_MEDIAN | mean arterial blood pressure | median |
| 10 | BP_sys_MEDIAN | systolic arterial blood pressure | median |
| 11 | INR_MAX | international normalized ratio of thrombin time | maximum |
| 12 | K_MAX | potassium level (serum) | maximum |
| 13 | CREA_MAX | creatinine level (serum) | maximum |
| 14 | LAC_MAX | lactate level (point-of-care testing) | maximum |
| 15 | NA_MIN | sodium level (serum or point-of-care testing) | minimum |
| 16 | SpO2_MIN | peripheral oxygen saturation | minimum |
| 17 | PLT_MIN | platelet count (blood) | minimum |
| 18 | WBC_MIN | white blood cells count (blood) | minimum |
| 19 | APTT_MAX | activated partial thromboplastin time | maximum |
| 20 | pCO2_MAX | partial pressure of carbon dioxide (arterial blood gas) | maximum |
| 21 | pO2_MIN | partial pressure of oxygen (arterial blood gas) | minimum |
| 22 | sO2_MIN | oxygen saturation (arterial blood gas) | minimum |
| 23 | HCO3- | bicarbonate (blood or point-of-care testing) | maximum |
| 24 | BP_dia_MEDIAN | diastolic arterial blood pressure | median |
| 25 | Quick_MIN | quick-value | minimum |

**Supplementary Table 2:** Clinical features that were used for machine learning.

| # | Abbreviation | Description | Location parameter |
|---|---|---|---|
| 1 | ALP_MAX | alkaline phosphatase level (serum) | maximum |
| 2 | BE_MEDIAN | base excess (arterial blood gas) | median |
| 3 | BR_total_MAX | total bilirubin level (serum) | maximum |
| 4 | CK_MAX | creatine kinase level (serum) | maximum |
| 5 | CRP_MAX | C-reactive protein level (serum) | maximum |
| 6 | CA_ionized_MIN | ionized calcium level (serum or point-of-care testing) | minimum |
| 7 | TC_MAX | total cholesterol level (serum) | maximum |
| 8 | CL_MIN | chlorine level (serum of point-of-care testing) | minimum |
| 9 | PROT_total_MIN | total protein level (serum) | minimum |
| 10 | FiO2_MAX | fraction of inspired oxygen | maximum |
| 11 | FIBR_MIN | fibrinogen level (serum) | minimum |
| 12 | GGT_MAX | gamma-glutamyltransferase level (serum) | maximum |
| 13 | AST_MAX | aspartate transaminase level (serum) | maximum |
| 14 | ALT_MAX | alanine transaminase level (serum) | maximum |
| 15 | HCO3-_MAX | bicarbonate (blood or point-of-care testing) | maximum |
| 16 | HR_MEDIAN | heart rate | median |
| 17 | UREA_N_MAX | urea nitrogen level (serum) | maximum |
| 18 | HGB_MIN | hemoglobin level (blood) | minimum |
| 19 | BP_dia_MEDIAN | diastolic arterial blood pressure | median |
| 20 | BP_mean_MEDIAN | mean arterial blood pressure | median |
| 21 | BP_sys_MEDIAN | systolic arterial blood pressure | median |
| 22 | INR_MAX | international normalized ratio of thrombin time | maximum |
| 23 | K_MAX | potassium level (serum) | maximum |
| 24 | CREA_MAX | creatinine level (serum) | maximum |
| 25 | LDH_MAX | lactate dehydrogenase level (serum) | maximum |
| 26 | LAC_MAX | lactate level (point-of-care testing) | maximum |
| 27 | LIP_MAX | lipase level (serum) | maximum |
| 28 | NA_MIN | sodium level (serum) or point-of-care testing) | minimum |
| 29 | PCT_MAX | procalcitonin level (serum) | maximum |
| 30 | PH_MIN | phosphate level (serum) | minimum |
| 31 | Quick_MIN | quick-value | minimum |
| 32 | SpO2_MIN | peripheral oxygen saturation | minimum |
| 33 | TEMP_MAX | body temperature | maximum |
| 34 | PLT_MIN | platelet count (blood) | minimum |
| 35 | TG_MAX | trigylceride level (serum) | maximum |
| 36 | WBC_MIN | white blood cells count (blood) | minimum |
| 37 | APTT_MAX | activated partial thromboplastin time | maximum |
| 38 | pCO2_MAX | partial pressure of carbon dioxide (arterial blood gas) | maximum |
| 39 | pH_MIN | pH (potential of hydrogen, arterial blood gas) | minimum |
| 40 | pO2_MIN | partial pressure of oxygen (arterial blood gas) | minimum |
| 41 | sO2_MIN | oxygen saturation (arterial blood gas) | minimum |

**Supplementary Table 3:** Correlating feature pairs in the clinical dataset as evaluated by the Pearson Correlation Coefficient.

| Feature 1 | Feature 2 | Pearson Correlation Coefficient |
|---|---|---|
| BE_MEDIAN* | HCO3-_MAX | 0.913 |
| CL_MIN | NA_MIN* | 0.733 |
| AST_MAX* | ALT_MAX* | 0.819 |
| AST_MAX* | LDH_MAX | 0.922 |
| ALT_MAX* | LDH_MAX | 0.800 |
| BP_dia_MEDIAN | BP_mean_MEDIAN* | 0.868 |
| INR_MAX* | LAC_MAX | 0.709 |
| INR_MAX* | Quick_MIN | -0.808 |
| pO2_MIN* | sO2_MIN | 0.728 |

*Features marked with a star (*) remained in the dataset, the other features were dropped.*

**Supplementary Table 4:** Hyperparameters for the random forest models for the prediction with clinical and proteomics data.

| Hyperparameter | Chosen value |
|---|---|
| n_estimators | 100 |
| min_samples_leaf | 5 |
| min_samples_split | 5 |
| max_depth | 10 |
| Class_weight | "balanced" |

**Supplementary Table 6:** Individual patients and their assignment to cluster A, B and C across the studied time points.

| Patient Identifier | SOFA | 30-day survival | Day 1 | Day 4 | Day 7 | Day 10 |
|---|---|---|---|---|---|---|
| KH-010 | 5 | Survival | A | A | | |
| KH-013 | 6 | Exitus | A | A | A | A |
| KH-019 | 2 | Survival | A | A | A | A |
| KH-022 | 6 | Exitus | A | A | B | B |
| KH-026 | 9 | Survival | A | A | A | A |
| KH-027 | 7 | Exitus | A | B | B | B |
| KH-028 | 9 | Exitus | A | A | B | B |
| KH-045 | 9 | Survival | A | A | B | |
| KH-046 | 13 | Survival | A | A | A | A |
| KH-047 | 12 | Survival | A | A | A | A |
| KKB-008 | 11 | Exitus | A | | | |
| KKB-009 | 13 | Exitus | A | A | C | B |
| KKB-011 | 5 | Survival | A | A | A | |
| KKB-013 | 7 | Exitus | A | A | A | A |
| KKB-014 | 4 | Survival | A | A | | |
| KKB-019 | 14 | Exitus | A | A | A | B |
| KKB-020 | 11 | Survival | A | | | |
| KKB-021 | 5 | Survival | A | A | A | A |
| KKB-023 | 2 | Exitus | A | A | A | A |
| KKB-027 | 11 | Exitus | A | A | A | A |
| KKB-029 | 6 | Survival | A | A | A | A |
| KKB-031 | 8 | Exitus | A | | | |
| KKB-035 | 7 | Survival | A | A | A | A |
| KKB-036 | 8 | Survival | A | A | A | A |
| KKB-037 | 8 | Exitus | A | A | A | |
| KKB-038 | 10 | Survival | A | A | A | A |
| KKB-041 | 7 | Survival | A | A | | |
| KKB-044 | 4 | Survival | A | A | A | A |
| KKB-045 | 4 | Exitus | A | A | A | |
| KKB-046 | 8 | Survival | A | | | A |
| KKB-049 | 5 | Survival | A | | | |
| KKB-050 | 9 | Survival | A | A | A | A |
| KKB-051 | 3 | Exitus | A | A | | A |
| KKB-054 | 13 | Exitus | A | A | A | A |
| KKB-055 | 11 | Exitus | A | A | A | B |
| KKB-061 | 3 | Survival | A | A | B | A |
| KKB-063 | 3 | Survival | A | | | |
| KKB-067 | 6 | Survival | A | A | A | A |
| KKB-070 | 8 | Exitus | A | A | C | |
| KKB-073 | 11 | Exitus | A | A | A | B |
| KKB-074 | 8 | Survival | A | A | A | |
| KKB-075 | 4 | Survival | A | A | B | A |
| KKB-077 | 17 | Exitus | A | A | A | A |
| KKB-078 | 4 | Survival | A | | | |
| KKB-079 | 5 | Survival | A | A | A | A |
| KKB-080 | 7 | Survival | A | | | |
| KKB-081 | 16 | Survival | A | A | A | A |
| KKB-083 | 4 | Survival | A | A | | |
| KKB-084 | 6 | Survival | A | | | |
| KKB-086 | 0 | Survival | A | | | |
| KKB-089 | 5 | Survival | A | | | |

| | | | | | | |
|---|---|---|---|---|---|---|
| KKB-099 | 9 | Survival | A | A | A | |
| KKB-103 | 5 | Exitus | A | A | | |
| KKB-104 | 10 | Exitus | A | A | A | |
| KKB-105 | 4 | Survival | A | | | |
| KKB-107 | 5 | Survival | A | A | A | A |
| KKB-108 | 11 | Survival | A | A | | |
| KKB-112 | 11 | Exitus | A | A | | |
| KKB-113 | 6 | Survival | A | A | A | |
| KKB-114 | 16 | Exitus | A | | | |
| KKB-123 | 5 | Survival | A | A | A | |
| KKB-124 | 13 | Exitus | A | A | | |
| KKB-136 | 6 | Exitus | A | | | |
| KKB-137 | 9 | Survival | A | A | | |
| KKB-138 | 6 | Exitus | A | | | |
| KKB-148 | 2 | Survival | A | | | |
| KKB-149 | 4 | Exitus | A | | | |
| KKB-151 | 7 | Survival | A | A | A | A |
| KKB-152 | 4 | Survival | A | | | |
| KKB-153 | 6 | Survival | A | | | |
| KKB-154 | 6 | Survival | A | | | |
| KKB-155 | 4 | Survival | A | | | |
| KKB-156 | 0 | Survival | A | | | |
| KKB-157 | 10 | Exitus | A | A | A | A |
| KKB-162 | 11 | Exitus | A | | | |
| KKB-167 | 15 | Exitus | A | A | B | A |
| KKB-168 | 12 | Exitus | A | | | |
| KKB-178 | 9 | Survival | A | A | A | A |
| KKB-182 | 8 | Survival | A | A | A | A |
| KKB-183 | 13 | Exitus | A | | | |
| KKB-198 | 15 | Exitus | A | A | B | B |
| KKB-202 | 12 | Exitus | A | A | B | B |
| KKB-209 | 3 | Survival | A | | | |
| KKB-210 | 9 | Exitus | A | | | |
| KKB-219 | 9 | Survival | A | A | A | A |
| KKB-220 | 7 | Exitus | A | | | |
| KKB-222 | 3 | Survival | A | | | |
| KKB-223 | 2 | Survival | A | | | |
| KKB-229 | 5 | Exitus | A | A | A | A |
| KKB-230 | 6 | Survival | A | | | |
| KKB-231 | 14 | Survival | A | A | A | A |
| KKB-233 | 9 | Exitus | A | A | B | B |
| KKB-237 | 8 | Exitus | A | A | A | A |
| KKB-238 | 11 | Exitus | A | A | A | A |
| KKB-242 | 8 | Exitus | A | B | A | A |
| KKB-244 | 5 | Exitus | A | A | A | A |
| KKB-245 | 8 | Exitus | A | | | |
| KKB-248 | 5 | Survival | A | A | B | B |
| KKB-249 | 1 | Survival | A | | | |
| KKB-250 | 2 | Survival | A | A | A | A |
| KKB-251 | 3 | Survival | A | A | A | |
| KKB-253 | 12 | Survival | A | A | A | B |
| KKB-254 | 2 | Exitus | A | A | A | A |
| KKB-255 | 5 | Survival | A | A | | |
| KKB-256 | 3 | Survival | A | A | A | |
| KKB-257 | 8 | Exitus | A | A | A | A |

| | | | | | | |
|---|---|---|---|---|---|---|
| KKB-258 | 3 | Survival | A | A | | |
| KKB-259 | 3 | Survival | A | A | | |
| KKB-260 | 3 | Survival | A | | | |
| KKB-261 | 12 | Survival | A | A | A | A |
| KKB-262 | 4 | Survival | A | A | A | |
| KKB-264 | 16 | Survival | A | A | A | A |
| KKB-265 | 10 | Survival | A | A | A | A |
| KKB-267 | 13 | Exitus | A | A | A | A |
| KKB-268 | 12 | Exitus | A | A | A | |
| KKB-269 | 5 | Survival | A | A | | |
| KKB-270 | 5 | Survival | A | A | | |
| KKB-271 | 7 | Survival | A | A | A | A |
| KKB-272 | 9 | Survival | A | A | | |
| KKB-274 | 9 | Survival | A | A | A | A |
| KKB-275 | 7 | Survival | A | A | A | A |
| KKB-276 | 2 | Survival | A | | | |
| KKB-277 | 2 | Survival | A | A | A | A |
| KKB-278 | 6 | Exitus | A | A | A | |
| KKB-279 | 9 | Survival | A | A | A | B |
| KKB-280 | 4 | Exitus | A | A | B | C |
| KKB-282 | 10 | Exitus | A | A | A | B |
| KKB-283 | 6 | Survival | A | A | | |
| KKB-284 | 14 | Survival | A | A | A | A |
| KKB-285 | 11 | Exitus | A | A | B | A |
| KKB-286 | 2 | Survival | A | | | |
| KKB-289 | 1 | Survival | A | | | |
| KKB-294 | 10 | Exitus | A | A | | |
| KKB-295 | 4 | Survival | A | A | A | B |
| KKB-296 | 3 | Exitus | A | A | A | A |
| KKB-297 | 3 | Survival | A | A | A | A |
| KKB-298 | 6 | Survival | A | A | | |
| KKB-299 | 8 | Exitus | A | A | A | A |
| KKB-300 | 11 | Survival | A | A | A | A |
| KKB-301 | 2 | Survival | A | | | |
| KKB-305 | 5 | Survival | A | | | |
| KKB-306 | 11 | Exitus | A | A | A | |
| KKB-307 | 13 | Survival | A | A | B | A |
| KKB-308 | 1 | Survival | A | | | |
| KKB-310 | 9 | Survival | A | B | B | B |
| KKB-312 | 6 | Exitus | A | B | B | A |
| KKB-317 | 8 | Survival | A | A | A | A |
| KKB-320 | 10 | Exitus | A | | | |
| KKB-321 | 8 | Survival | A | A | A | |
| KKB-322 | 5 | Survival | A | A | A | A |
| KKB-324 | 7 | Survival | A | A | A | A |
| KKB-326 | 19 | Exitus | A | A | B | B |
| KKB-327 | 14 | Exitus | A | A | A | A |
| KKB-328 | 14 | Survival | A | A | B | A |
| KKB-330 | 11 | Exitus | A | A | A | |
| KKB-331 | 15 | Exitus | A | B | | |
| KKB-332 | 13 | Survival | A | A | A | A |
| KKB-336 | 2 | Survival | A | A | A | |
| KKB-338 | 0 | Survival | A | | | |
| KKB-343 | 8 | Survival | A | A | | |
| KKB-345 | 7 | Survival | A | A | A | |

| | | | | | | |
|---|---|---|---|---|---|---|
| KKB-346 | 2 | Survival | A | | | |
| KKB-348 | 9 | Survival | A | A | A | A |
| KKB-350 | 6 | Exitus | A | A | A | |
| KKB-351 | 9 | Exitus | A | A | A | |
| KKB-353 | 12 | Survival | A | A | A | A |
| KKB-356 | 1 | Exitus | A | | | |
| KKB-357 | 6 | Survival | A | A | A | A |
| KKB-361 | 4 | Survival | A | | | |
| KKB-362 | 9 | Exitus | A | A | B | B |
| KKB-364 | 8 | Survival | A | A | A | A |
| KKB-365 | 11 | Survival | A | A | A | |
| KKB-370 | 8 | Exitus | A | B | B | B |
| KKB-372 | 14 | Survival | A | A | | |
| KKM-002 | 10 | Survival | A | A | B | A |
| KKM-004 | 3 | Survival | A | | | |
| KKM-013 | 3 | Survival | A | A | A | A |
| KKM-014 | 9 | Survival | A | A | A | A |
| KKM-015 | 3 | Survival | A | A | A | A |
| KKM-016 | 2 | Survival | A | | | |
| KKM-017 | 5 | Survival | A | | | |
| KKM-022 | 9 | Survival | A | A | A | |
| KKM-023 | 15 | Exitus | A | A | | |
| KKM-025 | 10 | Exitus | A | | | |
| KKM-026 | 8 | Survival | A | A | | |
| KKM-028 | 11 | Exitus | A | A | A | A |
| KKM-031 | 13 | Survival | A | A | A | A |
| KKM-032 | 16 | Survival | A | A | A | A |
| KKM-035 | 12 | Survival | A | A | | |
| KKM-041 | 2 | Exitus | A | A | A | |
| KKM-042 | 14 | Exitus | A | | | |
| KKM-043 | 6 | Exitus | A | A | A | A |
| KKM-044 | 3 | Survival | A | A | A | A |
| KKM-045 | 4 | Survival | A | | | |
| KKM-046 | 7 | Survival | A | | | |
| KKM-048 | 11 | Survival | A | A | | |
| KKM-050 | 12 | Survival | A | A | A | |
| KKM-051 | 12 | Exitus | A | A | A | A |
| KKM-055 | 14 | Survival | A | A | A | A |
| KKM-056 | 7 | Exitus | A | A | B | A |
| KKM-057 | 13 | Survival | A | A | A | A |
| KKM-059 | 5 | Survival | A | | | |
| KH-002 | 8 | Survival | B | B | A | B |
| KH-003 | 7 | Survival | B | A | A | A |
| KH-004 | 10 | Survival | B | A | A | |
| KH-005 | 8 | Exitus | B | A | A | A |
| KH-006 | 8 | Survival | B | A | A | |
| KH-007 | 8 | Survival | B | B | A | B |
| KH-008 | 6 | Exitus | B | B | B | A |
| KH-011 | 10 | Exitus | B | B | | |
| KH-012 | 11 | Exitus | B | B | B | B |
| KH-014 | 12 | Survival | B | B | A | A |
| KH-015 | 11 | Survival | B | A | B | A |
| KH-017 | 10 | Exitus | B | B | B | B |
| KH-018 | 4 | Survival | B | A | A | A |
| KH-020 | 2 | Survival | B | A | | |

| | | | | | | |
|---|---|---|---|---|---|---|
| KH-021 | 7 | Exitus | B | B | B | B |
| KH-023 | 11 | Survival | B | A | A | A |
| KH-024 | 7 | Survival | B | A | A | |
| KH-025 | 9 | Survival | B | B | A | A |
| KH-029 | 8 | Exitus | B | A | A | B |
| KH-030 | 6 | Exitus | B | B | A | |
| KH-031 | 9 | Survival | B | A | B | B |
| KH-032 | 5 | Exitus | B | | | |
| KH-034 | 12 | Exitus | B | B | B | A |
| KH-037 | 7 | Survival | B | B | | |
| KH-038 | 5 | Survival | B | B | | |
| KH-040 | 13 | Exitus | B | | | |
| KH-041 | 14 | Survival | B | B | B | A |
| KH-042 | 15 | Survival | B | A | A | A |
| KH-043 | 14 | Exitus | B | | | |
| KH-048 | 11 | Exitus | B | B | B | B |
| KH-049 | 12 | Survival | B | A | | |
| KH-050 | 13 | Survival | B | A | A | A |
| KH-051 | 11 | Exitus | B | A | B | B |
| KKB-010 | 3 | Survival | B | A | B | A |
| KKB-012 | 11 | Survival | B | A | A | A |
| KKB-017 | 11 | Exitus | B | C | | |
| KKB-022 | 15 | Exitus | B | B | B | B |
| KKB-024 | 7 | Survival | B | | | A |
| KKB-026 | 18 | Exitus | B | | | |
| KKB-032 | 5 | Exitus | B | | | |
| KKB-034 | 9 | Survival | B | A | B | B |
| KKB-042 | 15 | Exitus | B | | | |
| KKB-043 | 16 | Exitus | B | | | |
| KKB-048 | 11 | Survival | B | A | | |
| KKB-052 | 5 | Exitus | B | | | |
| KKB-056 | 8 | Exitus | B | B | B | C |
| KKB-057 | 5 | Survival | B | A | | |
| KKB-058 | 11 | Exitus | B | C | | |
| KKB-060 | 10 | Survival | B | | | |
| KKB-062 | 5 | Survival | B | B | B | |
| KKB-065 | 7 | Survival | B | B | | |
| KKB-066 | 14 | Exitus | B | C | | |
| KKB-069 | 19 | Exitus | B | B | B | |
| KKB-071 | 4 | Survival | B | | | |
| KKB-072 | 4 | Survival | B | | | |
| KKB-082 | 13 | Exitus | B | | | |
| KKB-085 | 6 | Exitus | B | A | | |
| KKB-087 | 6 | Survival | B | B | A | A |
| KKB-091 | 14 | Exitus | B | C | | |
| KKB-092 | 10 | Exitus | B | A | A | |
| KKB-093 | 4 | Survival | B | B | A | |
| KKB-094 | 8 | Survival | B | B | B | B |
| KKB-095 | 8 | Survival | B | A | A | |
| KKB-097 | 6 | Exitus | B | B | A | A |
| KKB-100 | 12 | Exitus | B | B | | |
| KKB-106 | 14 | Exitus | B | B | B | B |
| KKB-109 | 11 | Exitus | B | | | |
| KKB-110 | 7 | Survival | B | A | | |
| KKB-115 | 14 | Exitus | B | B | A | |

| | | | | | | |
|---|---|---|---|---|---|---|
| KKB-117 | 6 | Survival | B | A | | |
| KKB-127 | 1 | Survival | B | B | | |
| KKB-131 | 5 | Survival | B | | | |
| KKB-132 | 16 | Exitus | B | | | |
| KKB-143 | 16 | Exitus | B | | | |
| KKB-144 | 14 | Exitus | B | B | | |
| KKB-145 | 3 | Survival | B | A | A | |
| KKB-146 | 10 | Exitus | B | | | |
| KKB-147 | 14 | Exitus | B | A | | |
| KKB-159 | 12 | Survival | B | A | | |
| KKB-161 | 16 | Exitus | B | | | |
| KKB-170 | 10 | Exitus | B | | | |
| KKB-173 | 13 | Exitus | B | A | | |
| KKB-176 | 15 | Exitus | B | | | |
| KKB-177 | 13 | Exitus | B | C | | |
| KKB-181 | 4 | Survival | B | | | |
| KKB-184 | 11 | Exitus | B | A | A | B |
| KKB-190 | 14 | Exitus | B | B | C | |
| KKB-194 | 19 | Exitus | B | | | |
| KKB-197 | 17 | Exitus | B | B | B | |
| KKB-201 | 13 | Exitus | B | | | |
| KKB-215 | 10 | Survival | B | | | |
| KKB-232 | 11 | Exitus | B | A | | |
| KKB-235 | 14 | Exitus | B | C | | |
| KKB-236 | 15 | Exitus | B | A | A | A |
| KKB-239 | 18 | Exitus | B | B | A | B |
| KKB-240 | 14 | Exitus | B | A | B | B |
| KKB-243 | 10 | Exitus | B | | | |
| KKB-246 | 16 | Exitus | B | C | A | A |
| KKB-263 | 3 | Exitus | B | A | A | A |
| KKB-273 | 11 | Exitus | B | A | A | |
| KKB-287 | 10 | Survival | B | | | |
| KKB-290 | 7 | Exitus | B | B | | |
| KKB-293 | 16 | Survival | B | A | A | A |
| KKB-302 | 4 | Survival | B | | | |
| KKB-303 | 5 | Exitus | B | | | |
| KKB-304 | 13 | Survival | B | A | A | A |
| KKB-309 | 13 | Survival | B | A | A | B |
| KKB-311 | 4 | Survival | B | A | A | A |
| KKB-313 | 16 | Exitus | B | | | |
| KKB-315 | 15 | Exitus | B | | | |
| KKB-318 | 8 | Exitus | B | | | |
| KKB-319 | 19 | Exitus | B | B | B | |
| KKB-323 | 8 | Survival | B | A | | |
| KKB-325 | 8 | Survival | B | A | A | B |
| KKB-329 | 3 | Survival | B | | | |
| KKB-333 | 10 | Survival | B | B | A | B |
| KKB-334 | 2 | Survival | B | | | |
| KKB-335 | 9 | Exitus | B | B | B | B |
| KKB-337 | 6 | Exitus | B | | | |
| KKB-339 | 18 | Exitus | B | | | |
| KKB-340 | 2 | Survival | B | A | | |
| KKB-341 | 6 | Survival | B | | | |
| KKB-342 | 2 | Survival | B | | | |
| KKB-344 | 7 | Exitus | B | A | A | B |

| | | | | | | |
|---|---|---|---|---|---|---|
| KKB-347 | 7 | Exitus | B | A | | |
| KKB-349 | 6 | Survival | B | | | |
| KKB-355 | 15 | Survival | B | B | A | A |
| KKB-359 | 4 | Survival | B | A | | |
| KKB-363 | 9 | Survival | B | B | A | B |
| KKB-366 | 14 | Exitus | B | | | |
| KKB-367 | 12 | Exitus | B | A | B | |
| KKB-368 | 5 | Survival | B | | | |
| KKB-369 | 15 | Exitus | B | | | |
| KKB-371 | 5 | Survival | B | | | |
| KKM-001 | 11 | Survival | B | A | A | A |
| KKM-006 | 4 | Survival | B | A | | |
| KKM-007 | 5 | Survival | B | A | | |
| KKM-008 | 16 | Exitus | B | A | A | B |
| KKM-009 | 13 | Survival | B | B | A | A |
| KKM-010 | 8 | Survival | B | B | A | A |
| KKM-012 | 5 | Survival | B | A | | |
| KKM-021 | 10 | Survival | B | A | | |
| KKM-024 | 9 | Exitus | B | A | A | A |
| KKM-027 | 11 | Survival | B | A | A | |
| KKM-029 | 10 | Exitus | B | | | |
| KKM-030 | 13 | Survival | B | B | B | A |
| KKM-033 | 5 | Survival | B | B | | |
| KKM-034 | 10 | Survival | B | B | A | |
| KKM-037 | 13 | Exitus | B | | | |
| KKM-038 | 9 | Survival | B | B | A | B |
| KKM-039 | 10 | Exitus | B | B | | |
| KKM-040 | 12 | Exitus | B | | | |
| KKM-049 | 5 | Survival | B | | | |
| KKM-052 | 9 | Exitus | B | B | C | C |
| KKM-053 | 5 | Survival | B | A | A | A |
| KKM-054 | 2 | Survival | B | B | | |
| KH-009 | 12 | Exitus | C | B | | |
| KH-016 | 13 | Exitus | C | A | B | B |
| KH-033 | 9 | Exitus | C | B | A | A |
| KH-035 | 7 | Exitus | C | | | |
| KH-036 | 12 | Exitus | C | | | |
| KH-039 | 15 | Exitus | C | | | |
| KH-044 | 15 | Survival | C | A | A | A |
| KKB-015 | 14 | Exitus | C | A | B | B |
| KKB-018 | 20 | Exitus | C | | | |
| KKB-025 | 15 | Exitus | C | | | |
| KKB-028 | 13 | Exitus | C | | | |
| KKB-030 | 14 | Exitus | C | | | |
| KKB-064 | 13 | Exitus | C | B | B | B |
| KKB-076 | 17 | Exitus | C | | | |
| KKB-088 | 17 | Exitus | C | | | |
| KKB-134 | 16 | Exitus | C | | | |
| KKB-160 | 19 | Exitus | C | | | |
| KKB-169 | 14 | Exitus | C | | | |
| KKB-187 | 18 | Exitus | C | | | |
| KKB-189 | 19 | Exitus | C | | | |
| KKB-200 | 16 | Exitus | C | | | |
| KKB-316 | 14 | Exitus | C | | | |
| KKB-354 | 18 | Exitus | C | | | |

| | | | | | | |
|---|---|---|---|---|---|---|
| KKM-011 | 9 | Exitus | C | B | B | B |
| KKM-047 | 21 | Exitus | C | B | B | B |
| KKM-058 | 15 | Exitus | C | | | |

**Supplementary Table 7:** Significantly differentially abundant proteins as determined by plasma proteomics.

### Day 1 - Cluster A vs. Cluster B

| Protein | Gene | Description | Patients cluster A | Patients cluster B | ANOVA $p_{FDR}$-value | Posthoc p-value | Ratio of means B / A |
|---|---|---|---|---|---|---|---|
| P61769 | B2M | Beta-2-microglobulin | 146 | 110 | 7.73E-09 | 1.08E-10 | 1.78 |
| P00746 | CFD | Complement factor D | 146 | 110 | 1.43E-08 | 7.50E-10 | 1.68 |
| P01034 | CST3 | Cystatin-C | 144 | 110 | 1.27E-05 | 6.46E-07 | 1.33 |
| P61626 | LYZ | Lysozyme C | 140 | 108 | 1.90E-05 | 1.16E-06 | 1.44 |
| P36222 | CHI3L1 | Chitinase-3-like protein 1 | 83 | 92 | 1.04E-05 | 1.34E-06 | 2.06 |
| O43286 | B4GALT5 | Beta-1,4-galactosyl-transferase 5 | 142 | 105 | 5.48E-06 | 3.73E-06 | 1.42 |
| P04275 | VWF | von Willebrand factor | 146 | 110 | 1.16E-07 | 4.64E-06 | 1.45 |
| P02760 | AMBP | Protein AMBP | 146 | 110 | 1.19E-04 | 1.10E-05 | 1.25 |
| P80188 | LCN2 | Neutrophil gelatinase-associated lipocalin | 67 | 89 | 2.15E-04 | 3.03E-05 | 1.48 |
| P41222 | PTGDS | Prostaglandin-H2 D-isomerase | 86 | 87 | 1.44E-03 | 1.77E-04 | 1.35 |
| P35858 | IGFALS | Insulin-like growth factor-binding protein complex acid labile subunit | 146 | 108 | 1.36E-03 | 2.88E-04 | 0.76 |
| P05154 | SERPINA5 | Plasma serine protease inhibitor | 145 | 105 | 1.04E-06 | 5.93E-04 | 0.77 |
| Q9Y279 | VSIG4 | V-set and immunoglobulin domain-containing protein 4 | 92 | 93 | 6.37E-04 | 6.84E-04 | 1.66 |
| P36955 | SERPINF1 | Pigment epithelium-derived factor | 146 | 110 | 4.93E-03 | 9.79E-04 | 1.16 |
| P13473 | LAMP2 | Lysosome-associated membrane glycoprotein 2 | 73 | 42 | 3.23E-06 | 1.10E-03 | 0.71 |
| Q12805 | EFEMP1 | EGF-containing fibulin-like extracellular matrix protein 1 | 141 | 109 | 2.23E-04 | 1.24E-03 | 1.22 |
| P05062 | ALDOB | Fructose-bisphosphate aldolase B | 107 | 93 | 8.32E-16 | 1.44E-03 | 2.62 |
| P04180 | LCAT | Phosphatidylcholine-sterol acyltransferase | 146 | 110 | 1.92E-16 | 1.48E-03 | 0.87 |
| P13671 | C6 | Complement component C6 | 146 | 110 | 5.57E-09 | 1.78E-03 | 0.89 |
| P49908 | SELENOP | Selenoprotein P | 146 | 110 | 9.19E-06 | 1.79E-03 | 0.84 |

| | | | | | | | |
|---|---|---|---|---|---|---|---|
| P02753 | RBP4 | Retinol-binding protein 4 | 146 | 110 | 8.39E-03 | 2.46E-03 | 1.32 |
| P22692 | IGFBP4 | Insulin-like growth factor-binding protein 4 | 29 | 23 | 1.47E-02 | 3.04E-03 | 1.69 |
| P10909 | CLU | Clusterin | 146 | 110 | 2.23E-06 | 3.82E-03 | 0.89 |
| P12259 | F5 | Coagulation factor V | 146 | 110 | 1.39E-19 | 7.81E-03 | 0.88 |
| P18065 | IGFBP2 | Insulin-like growth factor-binding protein 2 | 135 | 105 | 4.56E-03 | 8.22E-03 | 1.44 |
| P0DJI9 | SAA2 | Serum amyloid A-2 protein | 146 | 110 | 5.07E-04 | 8.63E-03 | 1.53 |
| P04070 | PROC | Vitamin K-dependent protein C | 140 | 103 | 2.06E-19 | 9.77E-03 | 0.87 |
| P25311 | AZGP1 | Zinc-alpha-2-glycoprotein | 146 | 110 | 3.25E-02 | 1.32E-02 | 1.13 |
| P06727 | APOA4 | Apolipoprotein A-IV | 146 | 110 | 1.80E-02 | 1.43E-02 | 1.32 |
| P07339 | CTSD | Cathepsin D | 104 | 95 | 3.13E-03 | 1.81E-02 | 1.24 |
| P01024 | C3 | Complement C3 | 146 | 110 | 9.95E-14 | 1.87E-02 | 0.90 |
| P02741 | CRP | C-reactive protein | 146 | 110 | 3.98E-02 | 1.93E-02 | 1.37 |
| P37837 | TALDO1 | Transaldolase | 86 | 80 | 3.40E-13 | 2.14E-02 | 1.52 |
| P29622 | SERPINA4 | Kallistatin | 146 | 110 | 1.24E-03 | 2.31E-02 | 0.87 |
| P48740 | MASP1 | Mannan-binding lectin serine protease 1 | 146 | 109 | 1.59E-04 | 2.40E-02 | 0.89 |
| P07358 | C8B | Complement component C8 beta chain | 146 | 110 | 1.79E-04 | 2.58E-02 | 0.91 |
| P19320 | VCAM1 | Vascular cell adhesion protein 1 | 112 | 99 | 3.02E-02 | 2.98E-02 | 1.18 |
| P07237 | P4HB | Protein disulfide-isomerase | 82 | 69 | 5.70E-06 | 4.07E-02 | 1.19 |
| P02656 | APOC3 | Apolipoprotein C-III | 146 | 110 | 1.34E-04 | 4.21E-02 | 1.29 |
| P02748 | C9 | Complement component C9 | 146 | 110 | 2.72E-06 | 4.63E-02 | 0.92 |

**Day 1 - Cluster A vs. Cluster C**

| Protein | Gene | Description | Patients cluster A | Patients cluster C | ANOVA $p_{FDR}$-value | Posthoc p-value | Ratio of means C / A |
|---|---|---|---|---|---|---|---|
| P05062 | ALDOB | Fructose-bisphosphate aldolase B | 107 | 20 | 8.32E-16 | 0.00E+00 | 10.08 |
| P07900 | HSP90AA1 | Heat shock protein HSP 90-alpha | 109 | 20 | 7.81E-14 | 0.00E+00 | 4.14 |
| P62805 | H4C16 | Histone H4 | 85 | 20 | 1.30E-12 | 2.42E-14 | 9.30 |
| P0C0S8 | H2AC17 | Histone H2A type 1 | 112 | 20 | 3.14E-17 | 2.50E-14 | 15.99 |
| P00338 | LDHA | L-lactate dehydrogenase A chain | 120 | 20 | 4.41E-26 | 2.85E-14 | 11.62 |
| O60814 | H2BC12 | Histone H2B type 1-K | 124 | 20 | 1.07E-19 | 3.14E-14 | 14.68 |
| P04406 | GAPDH | Glyceraldehyde-3-phosphate dehydrogenase | 112 | 20 | 1.92E-16 | 3.35E-14 | 8.12 |
| P37837 | TALDO1 | Transaldolase | 86 | 20 | 3.40E-13 | 6.69E-14 | 4.26 |
| P78417 | GSTO1 | Glutathione S-transferase omega-1 | 117 | 20 | 3.69E-13 | 6.85E-14 | 4.70 |
| P29401 | TKT | Transketolase | 111 | 20 | 8.61E-12 | 1.18E-13 | 3.51 |
| Q14520 | HABP2 | Hyaluronan-binding protein 2 | 143 | 15 | 3.23E-20 | 1.38E-13 | 0.29 |
| P04070 | PROC | Vitamin K-dependent protein C | 140 | 15 | 2.06E-19 | 2.14E-13 | 0.29 |
| P12259 | F5 | Coagulation factor V | 146 | 20 | 1.39E-19 | 2.45E-13 | 0.42 |
| P60709 | ACTB | Actin, cytoplasmic 1 | 146 | 20 | 1.87E-15 | 2.50E-13 | 5.90 |
| P04004 | VTN | Vitronectin | 146 | 20 | 1.14E-17 | 2.50E-13 | 0.47 |
| P04180 | LCAT | Phosphatidylcholine-sterol acyltransferase | 146 | 20 | 1.92E-16 | 2.51E-13 | 0.47 |
| P01024 | C3 | Complement C3 | 146 | 20 | 9.95E-14 | 2.52E-13 | 0.59 |
| P05160 | F13B | Coagulation factor XIII B chain | 143 | 19 | 4.48E-13 | 2.65E-13 | 0.47 |
| P03951 | F11 | Coagulation factor XI | 145 | 20 | 9.41E-09 | 1.61E-10 | 0.60 |
| P68431 | H3C12 | Histone H3.1 | 55 | 20 | 1.10E-08 | 3.38E-10 | 8.06 |
| P13671 | C6 | Complement component C6 | 146 | 20 | 5.57E-09 | 4.15E-10 | 0.64 |
| Q16851 | UGP2 | UTP--glucose-1-phosphate uridylyltransferase | 73 | 20 | 1.82E-08 | 5.80E-10 | 6.73 |

| | | | | | | | |
|---|---|---|---|---|---|---|---|
| P21549 | AGXT | Alanine--glyoxylate aminotransferase | 81 | 18 | 3.43E-08 | 6.46E-10 | 4.86 |
| P06733 | ENO1 | Alpha-enolase | 82 | 20 | 7.01E-08 | 1.79E-09 | 4.63 |
| P11142 | HSPA8 | Heat shock cognate 71 kDa protein | 87 | 20 | 5.39E-08 | 2.64E-09 | 3.49 |
| P0C0L5 | C4B_2 | Complement C4-B | 146 | 20 | 1.45E-07 | 3.69E-09 | 0.59 |
| P20851 | C4BPB | C4b-binding protein beta chain | 139 | 20 | 2.08E-07 | 4.73E-09 | 0.55 |
| Q96KN2 | CNDP1 | Beta-Ala-His dipeptidase | 114 | 16 | 2.31E-07 | 5.87E-09 | 0.45 |
| P26927 | MST1 | Hepatocyte growth factor-like protein | 138 | 16 | 2.46E-07 | 6.33E-09 | 0.56 |
| P04040 | CAT | Catalase | 112 | 20 | 8.74E-07 | 2.33E-08 | 2.25 |
| P04075 | ALDOA | Fructose-bisphosphate aldolase A | 122 | 20 | 8.98E-07 | 2.53E-08 | 2.45 |
| P43251 | BTD | Biotinidase | 146 | 20 | 1.01E-06 | 3.25E-08 | 0.66 |
| P06276 | BCHE | Cholinesterase | 109 | 14 | 1.20E-06 | 3.78E-08 | 0.57 |
| P08238 | HSP90AB1 | Heat shock protein HSP 90-beta | 68 | 20 | 8.98E-07 | 8.62E-08 | 3.30 |
| P17174 | GOT1 | Aspartate aminotransferase, cytoplasmic | 66 | 20 | 2.89E-06 | 1.11E-07 | 4.42 |
| P05156 | CFI | Complement factor I | 146 | 20 | 3.34E-06 | 1.21E-07 | 0.67 |
| P02748 | C9 | Complement component C9 | 146 | 20 | 2.72E-06 | 1.65E-07 | 0.68 |
| Q04756 | HGFAC | Hepatocyte growth factor activator | 136 | 15 | 4.12E-06 | 2.11E-07 | 0.55 |
| P11021 | HSPA5 | Endoplasmic reticulum chaperone BiP | 131 | 20 | 5.70E-06 | 2.29E-07 | 2.36 |
| P37802 | TAGLN2 | Transgelin-2 | 41 | 9 | 7.04E-06 | 3.17E-07 | 0.36 |
| Q9UNW1 | MINPP1 | Multiple inositol polyphosphate phosphatase 1 | 60 | 15 | 6.59E-06 | 3.18E-07 | 0.56 |
| P07237 | P4HB | Protein disulfide-isomerase | 82 | 18 | 5.70E-06 | 3.24E-07 | 2.69 |
| P63104 | YWHAZ | 14-3-3 protein zeta/delta | 99 | 20 | 7.67E-06 | 3.34E-07 | 1.98 |
| P10909 | CLU | Clusterin | 146 | 20 | 2.23E-06 | 5.12E-07 | 0.69 |
| P13796 | LCP1 | Plastin-2 | 145 | 20 | 8.21E-06 | 5.42E-07 | 1.77 |
| P08263 | GSTA1 | Glutathione S-transferase A1 | 65 | 19 | 9.17E-06 | 6.30E-07 | 7.77 |
| P04278 | SHBG | Sex hormone-binding globulin | 127 | 18 | 9.74E-06 | 6.48E-07 | 0.54 |

| | | | | | | | |
|---|---|---|---|---|---|---|---|
| Q13228 | SELENBP1 | Methanethiol oxidase | 73 | 20 | 9.17E-06 | 7.38E-07 | 1.99 |
| Q16610 | ECM1 | Extracellular matrix protein 1 | 116 | 15 | 2.37E-06 | 7.89E-07 | 0.66 |
| P32119 | PRDX2 | Peroxiredoxin-2 | 143 | 20 | 1.54E-05 | 9.42E-07 | 2.32 |
| P05452 | CLEC3B | Tetranectin | 125 | 17 | 1.78E-05 | 9.82E-07 | 0.56 |
| P05154 | SERPINA5 | Plasma serine protease inhibitor | 145 | 16 | 1.04E-06 | 9.98E-07 | 0.48 |
| P00915 | CA1 | Carbonic anhydrase 1 | 142 | 20 | 1.80E-05 | 1.05E-06 | 2.73 |
| P26038 | MSN | Moesin | 97 | 20 | 1.10E-05 | 1.24E-06 | 2.17 |
| P15259 | PGAM2 | Phosphoglycerate mutase 2 | 43 | 20 | 1.82E-05 | 1.49E-06 | 3.08 |
| Q9UK55 | SERPINA10 | Protein Z-dependent protease inhibitor | 146 | 20 | 2.38E-05 | 1.68E-06 | 0.63 |
| P13473 | LAMP2 | Lysosome-associated membrane glycoprotein 2 | 73 | 9 | 3.23E-06 | 2.15E-06 | 0.40 |
| P04275 | VWF | von Willebrand factor | 146 | 20 | 1.16E-07 | 2.23E-06 | 2.26 |
| P55056 | APOC4 | Apolipoprotein C-IV | 132 | 11 | 3.96E-05 | 2.98E-06 | 0.40 |
| P68871 | HBB | Hemoglobin subunit beta | 146 | 20 | 5.27E-05 | 4.77E-06 | 2.71 |
| P02144 | MB | Myoglobin | 91 | 20 | 6.71E-05 | 5.20E-06 | 5.91 |
| P49908 | SELENOP | Selenoprotein P | 146 | 20 | 9.19E-06 | 6.92E-06 | 0.62 |
| P16930 | FAH | Fumarylacetoacetase | 39 | 20 | 1.08E-04 | 7.73E-06 | 3.55 |
| Q9NZP8 | C1RL | Complement C1r subcomponent-like protein | 146 | 19 | 1.15E-04 | 9.29E-06 | 0.73 |
| P05109 | S100A8 | Protein S100-A8 | 144 | 20 | 1.32E-04 | 9.58E-06 | 3.51 |
| O00187 | MASP2 | Mannan-binding lectin serine protease 2 | 145 | 19 | 1.27E-04 | 9.58E-06 | 0.59 |
| Q13790 | APOF | Apolipoprotein F | 131 | 14 | 7.93E-05 | 1.13E-05 | 0.49 |
| Q15848 | ADIPOQ | Adiponectin | 99 | 9 | 1.08E-04 | 1.20E-05 | 0.40 |
| P99999 | CYCS | Cytochrome c | 24 | 20 | 6.38E-05 | 1.29E-05 | 2.66 |
| P68032 | ACTC1 | Actin, alpha cardiac muscle 1 | 133 | 19 | 1.59E-04 | 1.34E-05 | 2.39 |
| P00742 | F10 | Coagulation factor X | 146 | 20 | 1.62E-04 | 1.85E-05 | 0.75 |
| P19971 | TYMP | Thymidine phosphorylase | 55 | 20 | 2.54E-04 | 2.96E-05 | 2.64 |

| | | | | | | | |
|---|---|---|---|---|---|---|---|
| P01602 | IGKV1-5 | Immunoglobulin kappa variable 1-5 | 123 | 16 | 4.09E-04 | 3.63E-05 | 0.58 |
| P07357 | C8A | Complement component C8 alpha chain | 146 | 20 | 2.14E-04 | 3.73E-05 | 0.76 |
| P48740 | MASP1 | Mannan-binding lectin serine protease 1 | 146 | 20 | 1.59E-04 | 4.69E-05 | 0.69 |
| P01833 | PIGR | Polymeric immunoglobulin receptor | 86 | 12 | 7.95E-05 | 5.32E-05 | 0.50 |
| P07358 | C8B | Complement component C8 beta chain | 146 | 20 | 1.79E-04 | 5.32E-05 | 0.74 |
| P69905 | HBA2 | Hemoglobin subunit alpha | 146 | 20 | 5.13E-04 | 5.33E-05 | 2.31 |
| A0A0A0MT36 | IGKV6D-21 | Immunoglobulin kappa variable 6D-21 | 136 | 18 | 2.99E-04 | 5.37E-05 | 0.48 |
| P00738 | HP | Haptoglobin | 146 | 20 | 6.02E-04 | 6.42E-05 | 0.61 |
| P00751 | CFB | Complement factor B | 146 | 20 | 6.37E-04 | 7.45E-05 | 0.75 |
| A0A0B4J1Y9 | IGHV3-72 | Immunoglobulin heavy variable 3-72 | 126 | 16 | 4.24E-04 | 7.92E-05 | 0.84 |
| P14151 | SELL | L-selectin | 112 | 17 | 4.52E-04 | 8.62E-05 | 0.57 |
| Q9BXR6 | CFHR5 | Complement factor H-related protein 5 | 145 | 18 | 4.24E-04 | 9.40E-05 | 0.59 |
| P68104 | EEF1A1 | Elongation factor 1-alpha 1 | 78 | 19 | 4.34E-04 | 1.19E-04 | 1.95 |
| P02790 | HPX | Hemopexin | 146 | 20 | 5.77E-04 | 1.37E-04 | 0.77 |
| P06702 | S100A9 | Protein S100-A9 | 145 | 20 | 1.44E-03 | 1.80E-04 | 3.22 |
| P19823 | ITIH2 | Inter-alpha-trypsin inhibitor heavy chain H2 | 146 | 20 | 1.58E-03 | 1.97E-04 | 0.77 |
| P02743 | APCS | Serum amyloid P-component | 146 | 20 | 1.03E-03 | 2.11E-04 | 0.70 |
| O75144 | ICOSLG | ICOS ligand | 67 | 10 | 1.34E-03 | 2.32E-04 | 0.48 |
| Q03154 | ACY1 | Aminoacylase-1 | 34 | 20 | 1.36E-03 | 2.51E-04 | 2.10 |
| P36980 | CFHR2 | Complement factor H-related protein 2 | 48 | 7 | 5.77E-04 | 2.55E-04 | 0.39 |
| P40925 | MDH1 | Malate dehydrogenase, cytoplasmic | 24 | 20 | 4.59E-04 | 2.91E-04 | 2.93 |
| P22891 | PROZ | Vitamin K-dependent protein Z | 119 | 15 | 1.66E-03 | 3.03E-04 | 0.48 |
| P04179 | SOD2 | Superoxide dismutase [Mn], mitochondrial | 81 | 20 | 6.87E-04 | 3.29E-04 | 2.87 |
| P01717 | IGLV3-25 | Immunoglobulin lambda variable 3-25 | 63 | 5 | 2.22E-03 | 3.42E-04 | 0.14 |
| Q96IY4 | CPB2 | Carboxypeptidase B2 | 146 | 20 | 1.36E-03 | 3.42E-04 | 0.77 |

| | | | | | | | |
|---|---|---|---|---|---|---|---|
| P01782 | IGHV3-9 | Immunoglobulin heavy variable 3-9 | 64 | 11 | 1.76E-03 | 3.61E-04 | 0.47 |
| Q9UGM5 | FETUB | Fetuin-B | 144 | 19 | 1.42E-03 | 3.76E-04 | 0.62 |
| O00151 | PDLIM1 | PDZ and LIM domain protein 1 | 27 | 9 | 2.63E-03 | 3.78E-04 | 0.44 |
| P28838 | LAP3 | Cytosol aminopeptidase | 37 | 18 | 1.88E-03 | 4.04E-04 | 2.33 |
| P00747 | PLG | Plasminogen | 146 | 20 | 1.61E-03 | 4.70E-04 | 0.78 |
| P02792 | FTL | Ferritin light chain | 32 | 17 | 8.68E-04 | 4.80E-04 | 4.57 |
| P01019 | AGT | Angiotensinogen | 146 | 20 | 2.62E-03 | 4.90E-04 | 1.40 |
| Q6EMK4 | VASN | Vasorin | 46 | 15 | 2.15E-04 | 5.40E-04 | 0.65 |
| P30101 | PDIA3 | Protein disulfide-isomerase A3 | 83 | 19 | 3.34E-03 | 5.91E-04 | 2.61 |
| Q06830 | PRDX1 | Peroxiredoxin-1 | 64 | 20 | 2.48E-03 | 6.10E-04 | 1.99 |
| P24158 | PRTN3 | Myeloblastin | 100 | 20 | 4.06E-03 | 6.21E-04 | 2.17 |
| P05362 | ICAM1 | Intercellular adhesion molecule 1 | 66 | 18 | 3.07E-03 | 6.80E-04 | 0.54 |
| P00488 | F13A1 | Coagulation factor XIII A chain | 142 | 20 | 4.48E-03 | 7.31E-04 | 0.72 |
| O75882 | ATRN | Attractin | 146 | 20 | 3.87E-03 | 7.44E-04 | 0.79 |
| P29622 | SERPINA4 | Kallistatin | 146 | 20 | 1.24E-03 | 7.73E-04 | 0.67 |
| P12955 | PEPD | Xaa-Pro dipeptidase | 99 | 20 | 4.70E-04 | 8.77E-04 | 1.77 |
| Q96PD5 | PGLYRP2 | N-acetylmuramoyl-L-alanine amidase | 146 | 20 | 4.82E-03 | 9.17E-04 | 0.75 |
| P09871 | C1S | Complement C1s subcomponent | 146 | 20 | 3.17E-03 | 1.06E-03 | 0.80 |
| P14618 | PKM | Pyruvate kinase PKM | 19 | 11 | 1.88E-03 | 1.14E-03 | 0.47 |
| P51884 | LUM | Lumican | 146 | 20 | 3.34E-03 | 1.14E-03 | 1.40 |
| P35542 | SAA4 | Serum amyloid A-4 protein | 146 | 20 | 6.30E-03 | 1.27E-03 | 0.68 |
| P31146 | CORO1A | Coronin-1A | 56 | 6 | 6.18E-03 | 1.30E-03 | 0.39 |
| P14625 | HSP90B1 | Endoplasmin | 82 | 20 | 4.06E-03 | 1.60E-03 | 1.92 |
| Q15942 | ZYX | Zyxin | 71 | 12 | 9.82E-03 | 1.85E-03 | 0.52 |
| P02656 | APOC3 | Apolipoprotein C-III | 146 | 20 | 1.34E-04 | 2.04E-03 | 0.63 |

| | | | | | | | |
|---|---|---|---|---|---|---|---|
| P00352 | ALDH1A1 | Aldehyde dehydrogenase 1A1 | 67 | 20 | 8.57E-03 | 2.08E-03 | 3.01 |
| P02774 | GC | Vitamin D-binding protein | 146 | 20 | 6.11E-03 | 2.56E-03 | 0.81 |
| P07339 | CTSD | Cathepsin D | 104 | 20 | 3.13E-03 | 2.60E-03 | 1.50 |
| P08519 | LPA | Apolipoprotein(a) | 131 | 17 | 1.16E-02 | 2.91E-03 | 0.57 |
| Q15582 | TGFBI | Transforming growth factor-beta-induced protein ig-h3 | 146 | 20 | 1.35E-02 | 2.94E-03 | 0.77 |
| A0A0B4J1U3 | IGLV1-36 | Immunoglobulin lambda variable 1-36 | 86 | 17 | 1.33E-02 | 3.04E-03 | 0.56 |
| P11226 | MBL2 | Mannose-binding protein C | 120 | 17 | 1.61E-02 | 3.24E-03 | 0.60 |
| O75019 | LILRA1 | Leukocyte immunoglobulin-like receptor subfamily A member 1 | 76 | 12 | 1.61E-02 | 3.55E-03 | 0.63 |
| P01008 | SERPINC1 | Antithrombin-III | 146 | 20 | 1.74E-02 | 3.65E-03 | 0.82 |
| P68363 | TUBA1B | Tubulin alpha-1B chain | 85 | 19 | 1.74E-02 | 3.71E-03 | 0.60 |
| P0DJI8 | SAA1 | Serum amyloid A-1 protein | 146 | 20 | 7.39E-04 | 3.74E-03 | 0.46 |
| A0A075B6S5 | IGKV1-27 | Immunoglobulin kappa variable 1-27 | 119 | 14 | 1.30E-02 | 4.16E-03 | 0.62 |
| P00325 | ADH1B | All-trans-retinol dehydrogenase [NAD(+)] ADH1B | 75 | 19 | 1.19E-02 | 4.36E-03 | 3.10 |
| Q9Y279 | VSIG4 | V-set and immunoglobulin domain-containing protein 4 | 92 | 19 | 6.37E-04 | 4.58E-03 | 1.95 |
| P25788 | PSMA3 | Proteasome subunit alpha type-3 | 27 | 19 | 1.55E-02 | 4.67E-03 | 2.34 |
| O95954 | FTCD | Formimidoyltransferase-cyclodeaminase | 9 | 17 | 4.25E-03 | 4.71E-03 | 3.69 |
| P07225 | PROS1 | Vitamin K-dependent protein S | 146 | 20 | 1.58E-02 | 5.08E-03 | 0.88 |
| P07148 | FABP1 | Fatty acid-binding protein, liver | 12 | 19 | 4.06E-03 | 5.61E-03 | 6.85 |
| P00740 | F9 | Coagulation factor IX | 146 | 20 | 2.35E-02 | 5.62E-03 | 0.82 |
| P00746 | CFD | Complement factor D | 146 | 20 | 1.43E-08 | 6.23E-03 | 1.48 |
| P42765 | ACAA2 | 3-ketoacyl-CoA thiolase, mitochondrial | 13 | 18 | 1.81E-02 | 6.44E-03 | 6.53 |
| P0DMV8 | HSPA1A | Heat shock 70 kDa protein 1A | 47 | 19 | 2.83E-02 | 6.69E-03 | 1.57 |
| P02655 | APOC2 | Apolipoprotein C-II | 146 | 19 | 1.47E-03 | 7.50E-03 | 0.57 |
| P36222 | CHI3L1 | Chitinase-3-like protein 1 | 83 | 19 | 1.04E-05 | 7.75E-03 | 1.80 |
| A0A0B4J1V2 | IGHV2-26 | Immunoglobulin heavy variable 2-26 | 119 | 14 | 2.26E-02 | 7.99E-03 | 0.57 |

| | | | | | | | |
|---|---|---|---|---|---|---|---|
| P04196 | HRG | Histidine-rich glycoprotein | 146 | 20 | 3.19E-02 | 8.55E-03 | 0.81 |
| P08697 | SERPINF2 | Alpha-2-antiplasmin | 146 | 20 | 2.11E-02 | 8.56E-03 | 0.86 |
| O75874 | IDH1 | Isocitrate dehydrogenase [NADP] cytoplasmic | 75 | 20 | 3.25E-02 | 8.86E-03 | 1.85 |
| P02654 | APOC1 | Apolipoprotein C-I | 146 | 20 | 2.21E-02 | 9.02E-03 | 0.74 |
| P61981 | YWHAG | 14-3-3 protein gamma | 7 | 10 | 2.72E-02 | 9.15E-03 | 1.79 |
| P27930 | IL1R2 | Interleukin-1 receptor type 2 | 11 | 6 | 3.43E-02 | 9.34E-03 | 2.40 |
| P06331 | IGHV4-34 | Immunoglobulin heavy variable 4-34 | 68 | 8 | 3.51E-02 | 9.34E-03 | 0.52 |
| P00739 | HPR | Haptoglobin-related protein | 145 | 20 | 1.61E-02 | 9.61E-03 | 0.73 |
| P0C0L4 | C4A | Complement C4-A | 146 | 20 | 1.29E-02 | 9.65E-03 | 0.62 |
| A0A0C4DH38 | IGHV5-51 | Immunoglobulin heavy variable 5-51 | 138 | 20 | 3.25E-02 | 9.69E-03 | 0.73 |
| P52209 | PGD | 6-phosphogluconate dehydrogenase, decarboxylating | 24 | 20 | 2.89E-02 | 9.77E-03 | 1.66 |
| Q7Z4W1 | DCXR | L-xylulose reductase | 16 | 19 | 2.16E-03 | 9.90E-03 | 2.32 |
| P33151 | CDH5 | Cadherin-5 | 75 | 10 | 3.24E-02 | 9.97E-03 | 0.43 |
| P33908 | MAN1A1 | Mannosyl-oligosaccharide 1,2-alpha-mannosidase IA | 137 | 20 | 5.87E-03 | 1.05E-02 | 0.89 |
| P09960 | LTA4H | Leukotriene A-4 hydrolase | 6 | 8 | 3.32E-02 | 1.05E-02 | 0.38 |
| Q08830 | FGL1 | Fibrinogen-like protein 1 | 133 | 19 | 3.14E-02 | 1.17E-02 | 0.58 |
| P14649 | MYL6B | Myosin light chain 6B | 41 | 10 | 4.21E-02 | 1.17E-02 | 0.64 |
| O43866 | CD5L | CD5 antigen-like | 145 | 20 | 4.54E-02 | 1.27E-02 | 1.60 |
| P30050 | RPL12 | 60S ribosomal protein L12 | 5 | 16 | 8.57E-03 | 1.51E-02 | 1.89 |
| P06744 | GPI | Glucose-6-phosphate isomerase | 35 | 19 | 1.33E-02 | 1.61E-02 | 1.41 |
| P00966 | ASS1 | Argininosuccinate synthase | 62 | 20 | 7.17E-03 | 1.83E-02 | 2.85 |
| P00505 | GOT2 | Aspartate aminotransferase, mitochondrial | 84 | 17 | 4.46E-02 | 1.86E-02 | 0.68 |
| P21333 | FLNA | Filamin-A | 108 | 20 | 3.53E-02 | 1.94E-02 | 0.63 |
| P18206 | VCL | Vinculin | 97 | 18 | 4.48E-02 | 2.09E-02 | 0.68 |
| P13639 | EEF2 | Elongation factor 2 | 11 | 18 | 2.32E-02 | 2.29E-02 | 2.03 |

| | | | | | | | |
|---|---|---|---|---|---|---|---|
| P80188 | LCN2 | Neutrophil gelatinase-associated lipocalin | 67 | 20 | 2.15E-04 | 2.37E-02 | 1.64 |
| P05543 | SERPINA7 | Thyroxine-binding globulin | 146 | 20 | 3.25E-02 | 2.45E-02 | 0.84 |
| P01871 | IGHM | Immunoglobulin heavy constant mu | 146 | 20 | 4.70E-02 | 2.52E-02 | 1.76 |
| P0DOX7 | | Immunoglobulin kappa light chain | 146 | 20 | 1.67E-02 | 2.57E-02 | 1.30 |
| P28066 | PSMA5 | Proteasome subunit alpha type-5 | 53 | 19 | 2.36E-02 | 2.60E-02 | 1.79 |
| O75369 | FLNB | Filamin-B | 21 | 11 | 3.24E-02 | 2.90E-02 | 0.44 |
| P06681 | C2 | Complement C2 | 146 | 20 | 4.70E-02 | 3.35E-02 | 0.87 |
| Q92954 | PRG4 | Proteoglycan 4 | 146 | 20 | 4.74E-02 | 3.61E-02 | 0.74 |
| P0DJI9 | SAA2 | Serum amyloid A-2 protein | 146 | 20 | 5.07E-04 | 3.95E-02 | 0.40 |

### Day 1 - Cluster B vs Cluster C

| Protein | Gene | Description | Patients cluster B | Patients cluster C | ANOVA $p_{FDR}$-value | Posthoc p-value | Ratio of means C / B |
|---|---|---|---|---|---|---|---|
| AP0C0S8 | H2AC17 | Histone H2A type 1 | 97 | 20 | 3.14E-17 | 2.55E-14 | 9.16 |
| P00338 | LDHA | L-lactate dehydrogenase A chain | 100 | 20 | 4.41E-26 | 2.90E-14 | 7.53 |
| P04406 | GAPDH | Glyceraldehyde-3-phosphate dehydrogenase | 91 | 20 | 1.92E-16 | 3.50E-14 | 6.05 |
| O60814 | H2BC12 | Histone H2B type 1-K | 98 | 20 | 1.07E-19 | 4.16E-14 | 9.81 |
| Q14520 | HABP2 | Hyaluronan-binding protein 2 | 105 | 15 | 3.23E-20 | 1.51E-13 | 0.33 |
| P04070 | PROC | Vitamin K-dependent protein C | 103 | 15 | 2.06E-19 | 2.20E-13 | 0.34 |
| P07900 | HSP90AA1 | Heat shock protein HSP 90-alpha | 85 | 20 | 7.81E-14 | 2.39E-13 | 3.55 |
| P12259 | F5 | Coagulation factor V | 110 | 20 | 1.39E-19 | 2.50E-13 | 0.48 |
| P04004 | VTN | Vitronectin | 110 | 20 | 1.14E-17 | 2.52E-13 | 0.50 |
| P60709 | ACTB | Actin, cytoplasmic 1 | 110 | 20 | 1.87E-15 | 2.96E-13 | 4.62 |
| P04180 | LCAT | Phosphatidylcholine-sterol acyltransferase | 110 | 20 | 1.92E-16 | 1.97E-12 | 0.54 |
| P78417 | GSTO1 | Glutathione S-transferase omega-1 | 87 | 20 | 3.69E-13 | 2.49E-12 | 3.22 |

| | | | | | | | |
|---|---|---|---|---|---|---|---|
| P05160 | F13B | Coagulation factor XIII B chain | 109 | 19 | 4.48E-13 | 2.59E-12 | 0.51 |
| P05062 | ALDOB | Fructose-bisphosphate aldolase B | 93 | 20 | 8.32E-16 | 6.58E-12 | 3.85 |
| P62805 | H4C16 | Histone H4 | 73 | 20 | 1.30E-12 | 2.42E-11 | 5.44 |
| P01024 | C3 | Complement C3 | 110 | 20 | 9.95E-14 | 2.89E-11 | 0.65 |
| P29401 | TKT | Transketolase | 84 | 20 | 8.61E-12 | 3.88E-11 | 2.75 |
| P37837 | TALDO1 | Transaldolase | 80 | 20 | 3.40E-13 | 9.07E-11 | 2.80 |
| P11142 | HSPA8 | Heat shock cognate 71 kDa protein | 72 | 20 | 5.39E-08 | 2.18E-08 | 2.86 |
| Q16851 | UGP2 | UTP--glucose-1-phosphate uridylyltransferase | 64 | 20 | 1.82E-08 | 3.00E-08 | 3.68 |
| P68431 | H3C12 | Histone H3.1 | 57 | 20 | 1.10E-08 | 3.97E-08 | 4.63 |
| P0C0L5 | C4B_2 | Complement C4-B | 110 | 20 | 1.45E-07 | 7.70E-08 | 0.60 |
| Q16610 | ECM1 | Extracellular matrix protein 1 | 77 | 15 | 2.37E-06 | 9.85E-08 | 0.62 |
| P26927 | MST1 | Hepatocyte growth factor-like protein | 102 | 16 | 2.46E-07 | 1.35E-07 | 0.61 |
| P06733 | ENO1 | Alpha-enolase | 74 | 20 | 7.01E-08 | 1.67E-07 | 1.95 |
| P03951 | F11 | Coagulation factor XI | 107 | 20 | 9.41E-09 | 1.72E-07 | 0.64 |
| P08238 | HSP90AB1 | Heat shock protein HSP 90-beta | 67 | 20 | 8.98E-07 | 2.79E-07 | 2.25 |
| P21549 | AGXT | Alanine--glyoxylate aminotransferase | 60 | 18 | 3.43E-08 | 3.92E-07 | 3.42 |
| Q04756 | HGFAC | Hepatocyte growth factor activator | 105 | 15 | 4.12E-06 | 6.93E-07 | 0.53 |
| P20851 | C4BPB | C4b-binding protein beta chain | 102 | 20 | 2.08E-07 | 7.09E-07 | 0.59 |
| P13796 | LCP1 | Plastin-2 | 110 | 20 | 8.21E-06 | 1.97E-06 | 1.76 |
| Q96KN2 | CNDP1 | Beta-Ala-His dipeptidase | 66 | 16 | 2.31E-07 | 2.77E-06 | 0.48 |
| P04278 | SHBG | Sex hormone-binding globulin | 98 | 18 | 9.74E-06 | 2.94E-06 | 0.54 |
| P26038 | MSN | Moesin | 72 | 20 | 1.10E-05 | 3.50E-06 | 2.11 |
| P05156 | CFI | Complement factor I | 110 | 20 | 3.34E-06 | 4.25E-06 | 0.69 |
| P04075 | ALDOA | Fructose-bisphosphate aldolase A | 96 | 20 | 8.98E-07 | 4.99E-06 | 1.98 |
| P55056 | APOC4 | Apolipoprotein C-IV | 107 | 11 | 3.96E-05 | 5.67E-06 | 0.41 |

| | | | | | | | |
|---|---|---|---|---|---|---|---|
| P01833 | PIGR | Polymeric immunoglobulin receptor | 49 | 12 | 7.95E-05 | 5.93E-06 | 0.45 |
| P04040 | CAT | Catalase | 90 | 20 | 8.74E-07 | 6.17E-06 | 1.88 |
| P06276 | BCHE | Cholinesterase | 58 | 14 | 1.20E-06 | 6.44E-06 | 0.61 |
| Q13228 | SELENBP1 | Methanethiol oxidase | 66 | 20 | 9.17E-06 | 6.85E-06 | 2.70 |
| P43251 | BTD | Biotinidase | 110 | 20 | 1.01E-06 | 7.04E-06 | 0.72 |
| P32119 | PRDX2 | Peroxiredoxin-2 | 110 | 20 | 1.54E-05 | 7.08E-06 | 2.16 |
| Q13790 | APOF | Apolipoprotein F | 102 | 14 | 7.93E-05 | 8.12E-06 | 0.43 |
| Q9UK55 | SERPINA10 | Protein Z-dependent protease inhibitor | 110 | 20 | 2.38E-05 | 9.30E-06 | 0.66 |
| P13671 | C6 | Complement component C6 | 110 | 20 | 5.57E-09 | 9.94E-06 | 0.71 |
| Q9UNW1 | MINPP1 | Multiple inositol polyphosphate phosphatase 1 | 59 | 15 | 6.59E-06 | 1.06E-05 | 0.61 |
| P00915 | CA1 | Carbonic anhydrase 1 | 108 | 20 | 1.80E-05 | 1.22E-05 | 2.63 |
| P68871 | HBB | Hemoglobin subunit beta | 110 | 20 | 5.27E-05 | 1.25E-05 | 2.59 |
| P17174 | GOT1 | Aspartate aminotransferase, cytoplasmic | 68 | 20 | 2.89E-06 | 1.28E-05 | 2.77 |
| P08263 | GSTA1 | Glutathione S-transferase A1 | 58 | 19 | 9.17E-06 | 1.28E-05 | 3.38 |
| P11021 | HSPA5 | Endoplasmic reticulum chaperone BiP | 104 | 20 | 5.70E-06 | 1.33E-05 | 2.11 |
| Q15848 | ADIPOQ | Adiponectin | 62 | 9 | 1.08E-04 | 1.43E-05 | 0.43 |
| P05452 | CLEC3B | Tetranectin | 88 | 17 | 1.78E-05 | 1.61E-05 | 0.58 |
| P02656 | APOC3 | Apolipoprotein C-III | 110 | 20 | 1.34E-04 | 1.77E-05 | 0.49 |
| Q6EMK4 | VASN | Vasorin | 59 | 15 | 2.15E-04 | 1.88E-05 | 0.58 |
| P37802 | TAGLN2 | Transgelin-2 | 29 | 9 | 7.04E-06 | 2.96E-05 | 0.42 |
| P07357 | C8A | Complement component C8 alpha chain | 110 | 20 | 2.14E-04 | 3.25E-05 | 0.75 |
| P12955 | PEPD | Xaa-Pro dipeptidase | 82 | 20 | 4.70E-04 | 4.51E-05 | 1.83 |
| A0A0A0MT36 | IGKV6D-21 | Immunoglobulin kappa variable 6D-21 | 104 | 18 | 2.99E-04 | 4.64E-05 | 0.46 |
| P63104 | YWHAZ | 14-3-3 protein zeta/delta | 77 | 20 | 7.67E-06 | 4.92E-05 | 1.69 |
| P36980 | CFHR2 | Complement factor H-related protein 2 | 39 | 7 | 5.77E-04 | 6.65E-05 | 0.35 |

| | | | | | | | |
|---|---|---|---|---|---|---|---|
| P15259 | PGAM2 | Phosphoglycerate mutase 2 | 49 | 20 | 1.82E-05 | 7.46E-05 | 2.15 |
| P14151 | SELL | L-selectin | 93 | 17 | 4.52E-04 | 7.97E-05 | 0.59 |
| P02790 | HPX | Hemopexin | 110 | 20 | 5.77E-04 | 8.82E-05 | 0.76 |
| P02748 | C9 | Complement component C9 | 110 | 20 | 2.72E-06 | 8.98E-05 | 0.73 |
| P68104 | EEF1A1 | Elongation factor 1-alpha 1 | 67 | 19 | 4.34E-04 | 9.66E-05 | 2.00 |
| P0DJI8 | SAA1 | Serum amyloid A-1 protein | 110 | 20 | 7.39E-04 | 1.04E-04 | 0.36 |
| P04179 | SOD2 | Superoxide dismutase [Mn], mitochondrial | 68 | 20 | 6.87E-04 | 1.21E-04 | 3.03 |
| P05109 | S100A8 | Protein S100-A8 | 107 | 20 | 1.32E-04 | 1.76E-04 | 3.15 |
| P99999 | CYCS | Cytochrome c | 28 | 20 | 6.38E-05 | 1.96E-04 | 2.06 |
| P02655 | APOC2 | Apolipoprotein C-II | 109 | 19 | 1.47E-03 | 2.58E-04 | 0.46 |
| O00187 | MASP2 | Mannan-binding lectin serine protease 2 | 110 | 19 | 1.27E-04 | 2.78E-04 | 0.63 |
| P0DJI9 | SAA2 | Serum amyloid A-2 protein | 110 | 20 | 5.07E-04 | 2.88E-04 | 0.26 |
| P69905 | HBA2 | Hemoglobin subunit alpha | 110 | 20 | 5.13E-04 | 2.93E-04 | 2.13 |
| P02792 | FTL | Ferritin light chain | 39 | 17 | 8.68E-04 | 2.99E-04 | 3.56 |
| P40925 | MDH1 | Malate dehydrogenase, cytoplasmic | 29 | 20 | 4.59E-04 | 3.14E-04 | 2.34 |
| P07237 | P4HB | Protein disulfide-isomerase | 69 | 18 | 5.70E-06 | 3.34E-04 | 2.25 |
| P19971 | TYMP | Thymidine phosphorylase | 52 | 20 | 2.54E-04 | 4.27E-04 | 1.90 |
| Q9NZP8 | C1RL | Complement C1r subcomponent-like protein | 110 | 19 | 1.15E-04 | 4.28E-04 | 0.78 |
| Q12805 | EFEMP1 | EGF-containing fibulin-like extracellular matrix protein 1 | 109 | 20 | 2.23E-04 | 4.41E-04 | 0.76 |
| O43286 | B4GALT5 | Beta-1,4-galactosyl-transferase 5 | 105 | 19 | 5.48E-06 | 4.45E-04 | 0.60 |
| P01602 | IGKV1-5 | Immunoglobulin kappa variable 1-5 | 95 | 16 | 4.09E-04 | 4.59E-04 | 0.67 |
| P01782 | IGHV3-9 | Immunoglobulin heavy variable 3-9 | 53 | 11 | 1.76E-03 | 5.09E-04 | 0.44 |
| Q7Z4W1 | DCXR | L-xylulose reductase | 23 | 19 | 2.16E-03 | 5.29E-04 | 2.46 |
| P68032 | ACTC1 | Actin, alpha cardiac muscle 1 | 91 | 19 | 1.59E-04 | 6.55E-04 | 2.32 |
| P01019 | AGT | Angiotensinogen | 110 | 20 | 2.62E-03 | 7.83E-04 | 1.37 |

| | | | | | | | |
|---|---|---|---|---|---|---|---|
| P14625 | HSP90B1 | Endoplasmin | 83 | 20 | 4.06E-03 | 9.55E-04 | 1.95 |
| P02144 | MB | Myoglobin | 88 | 20 | 6.71E-05 | 9.71E-04 | 2.39 |
| P33908 | MAN1A1 | Mannosyl-oligosaccharide 1,2-alpha-mannosidase IA | 109 | 20 | 5.87E-03 | 1.02E-03 | 0.82 |
| P14618 | PKM | Pyruvate kinase PKM | 13 | 11 | 1.88E-03 | 1.04E-03 | 0.39 |
| P10909 | CLU | Clusterin | 110 | 20 | 2.23E-06 | 1.18E-03 | 0.78 |
| P19823 | ITIH2 | Inter-alpha-trypsin inhibitor heavy chain H2 | 110 | 20 | 1.58E-03 | 1.22E-03 | 0.79 |
| P00738 | HP | Haptoglobin | 110 | 20 | 6.02E-04 | 1.26E-03 | 0.73 |
| O75882 | ATRN | Attractin | 109 | 20 | 3.87E-03 | 1.27E-03 | 0.78 |
| P00966 | ASS1 | Argininosuccinate synthase | 40 | 20 | 7.17E-03 | 1.28E-03 | 2.77 |
| P05362 | ICAM1 | Intercellular adhesion molecule 1 | 69 | 18 | 3.07E-03 | 1.40E-03 | 0.58 |
| P00742 | F10 | Coagulation factor X | 110 | 20 | 1.62E-04 | 1.47E-03 | 0.79 |
| Q06830 | PRDX1 | Peroxiredoxin-1 | 45 | 20 | 2.48E-03 | 1.62E-03 | 1.72 |
| Q96PD5 | PGLYRP2 | N-acetylmuramoyl-L-alanine amidase | 110 | 20 | 4.82E-03 | 1.85E-03 | 0.75 |
| P00751 | CFB | Complement factor B | 110 | 20 | 6.37E-04 | 1.89E-03 | 0.80 |
| Q00796 | SORD | Sorbitol dehydrogenase | 43 | 19 | 1.02E-02 | 1.97E-03 | 2.06 |
| P31146 | CORO1A | Coronin-1A | 27 | 6 | 6.18E-03 | 2.17E-03 | 0.38 |
| P07148 | FABP1 | Fatty acid-binding protein, liver | 28 | 19 | 4.06E-03 | 2.35E-03 | 2.77 |
| Q03154 | ACY1 | Aminoacylase-1 | 43 | 20 | 1.36E-03 | 2.39E-03 | 2.14 |
| P06702 | S100A9 | Protein S100-A9 | 108 | 20 | 1.44E-03 | 2.41E-03 | 3.09 |
| P30101 | PDIA3 | Protein disulfide-isomerase A3 | 65 | 19 | 3.34E-03 | 2.41E-03 | 2.19 |
| P00488 | F13A1 | Coagulation factor XIII A chain | 110 | 20 | 4.48E-03 | 2.61E-03 | 0.77 |
| P05154 | SERPINA5 | Plasma serine protease inhibitor | 105 | 16 | 1.04E-06 | 2.73E-03 | 0.62 |
| A0A075B6S5 | IGKV1-27 | Immunoglobulin kappa variable 1-27 | 87 | 14 | 1.30E-02 | 3.06E-03 | 0.61 |
| P08519 | LPA | Apolipoprotein(a) | 103 | 17 | 1.16E-02 | 3.39E-03 | 0.58 |
| P28838 | LAP3 | Cytosol aminopeptidase | 27 | 18 | 1.88E-03 | 3.51E-03 | 2.20 |

| | | | | | | | |
|---|---|---|---|---|---|---|---|
| P01717 | IGLV3-25 | Immunoglobulin lambda variable 3-25 | 28 | 5 | 2.22E-03 | 3.59E-03 | 0.20 |
| O95954 | FTCD | Formimidoyltransferase-cyclodeaminase | 21 | 17 | 4.25E-03 | 3.66E-03 | 2.75 |
| P0DOX7 | | Immunoglobulin kappa light chain | 110 | 20 | 1.67E-02 | 3.69E-03 | 1.39 |
| P06744 | GPI | Glucose-6-phosphate isomerase | 35 | 19 | 1.33E-02 | 3.73E-03 | 1.72 |
| P07225 | PROS1 | Vitamin K-dependent protein S | 110 | 20 | 1.58E-02 | 4.22E-03 | 0.87 |
| P00325 | ADH1B | All-trans-retinol dehydrogenase [NAD(+)] ADH1B | 55 | 19 | 1.19E-02 | 4.33E-03 | 1.90 |
| P16930 | FAH | Fumarylacetoacetase | 44 | 20 | 1.08E-04 | 5.28E-03 | 1.91 |
| P08697 | SERPINF2 | Alpha-2-antiplasmin | 110 | 20 | 2.11E-02 | 5.36E-03 | 0.84 |
| P22234 | PAICS | Bifunctional phosphoribosylaminoimidazole carboxylase/ phosphoribosyl-aminoimidazole succinocarboxamide synthetase | 5 | 9 | 1.71E-02 | 5.37E-03 | 2.09 |
| P02654 | APOC1 | Apolipoprotein C-I | 110 | 20 | 2.21E-02 | 5.66E-03 | 0.72 |
| P00352 | ALDH1A1 | Aldehyde dehydrogenase 1A1 | 56 | 20 | 8.57E-03 | 5.93E-03 | 2.69 |
| Q15582 | TGFBI | Transforming growth factor-beta-induced protein ig-h3 | 109 | 20 | 1.35E-02 | 5.95E-03 | 0.76 |
| A0A0B4J1Y9 | IGHV3-72 | Immunoglobulin heavy variable 3-72 | 89 | 16 | 4.24E-04 | 6.07E-03 | 0.94 |
| P28066 | PSMA5 | Proteasome subunit alpha type-5 | 44 | 19 | 2.36E-02 | 6.21E-03 | 1.87 |
| P13473 | LAMP2 | Lysosome-associated membrane glycoprotein 2 | 42 | 9 | 3.23E-06 | 6.73E-03 | 0.57 |
| P24158 | PRTN3 | Myeloblastin | 86 | 20 | 4.06E-03 | 7.10E-03 | 1.80 |
| P30050 | RPL12 | 60S ribosomal protein L12 | 14 | 16 | 8.57E-03 | 7.10E-03 | 2.01 |
| Q9BXR6 | CFHR5 | Complement factor H-related protein 5 | 109 | 18 | 4.24E-04 | 7.33E-03 | 0.66 |
| O00151 | PDLIM1 | PDZ and LIM domain protein 1 | 31 | 9 | 2.63E-03 | 7.48E-03 | 0.51 |
| O94760 | DDAH1 | N(G),N(G)-dimethylarginine dimethylamino-hydrolase 1 | 10 | 7 | 2.94E-02 | 7.60E-03 | 1.94 |
| O75144 | ICOSLG | ICOS ligand | 59 | 10 | 1.34E-03 | 7.96E-03 | 0.54 |
| P22891 | PROZ | Vitamin K-dependent protein Z | 84 | 15 | 1.66E-03 | 8.34E-03 | 0.57 |
| P18065 | IGFBP2 | Insulin-like growth factor-binding protein 2 | 105 | 20 | 4.56E-03 | 8.45E-03 | 0.53 |
| Q5TEC6 | H3-7 | Histone H3-7 | 10 | 16 | 7.14E-03 | 8.83E-03 | 1.76 |

| | | | | | | | |
|---|---|---|---|---|---|---|---|
| Q08830 | FGL1 | Fibrinogen-like protein 1 | 99 | 19 | 3.14E-02 | 9.62E-03 | 0.60 |
| P48740 | MASP1 | Mannan-binding lectin serine protease 1 | 109 | 20 | 1.59E-04 | 1.00E-02 | 0.78 |
| P49908 | SELENOP | Selenoprotein P | 110 | 20 | 9.19E-06 | 1.01E-02 | 0.73 |
| P21333 | FLNA | Filamin-A | 89 | 20 | 3.53E-02 | 1.03E-02 | 0.59 |
| P07358 | C8B | Complement component C8 beta chain | 110 | 20 | 1.79E-04 | 1.03E-02 | 0.81 |
| P02743 | APCS | Serum amyloid P-component | 110 | 20 | 1.03E-03 | 1.09E-02 | 0.78 |
| P33151 | CDH5 | Cadherin-5 | 58 | 10 | 3.24E-02 | 1.17E-02 | 0.46 |
| P00740 | F9 | Coagulation factor IX | 110 | 20 | 2.35E-02 | 1.18E-02 | 0.83 |
| P49720 | PSMB3 | Proteasome subunit beta type-3 | 8 | 11 | 4.20E-02 | 1.20E-02 | 0.28 |
| P23528 | CFL1 | Cofilin-1 | 83 | 19 | 4.66E-02 | 1.28E-02 | 1.68 |
| O75369 | FLNB | Filamin-B | 15 | 11 | 3.24E-02 | 1.30E-02 | 0.39 |
| P11226 | MBL2 | Mannose-binding protein C | 95 | 17 | 1.61E-02 | 1.30E-02 | 0.67 |
| P06681 | C2 | Complement C2 | 110 | 20 | 4.70E-02 | 1.30E-02 | 0.86 |
| P01008 | SERPINC1 | Antithrombin-III | 110 | 20 | 1.74E-02 | 1.31E-02 | 0.84 |
| P01871 | IGHM | Immunoglobulin heavy constant mu | 110 | 20 | 4.70E-02 | 1.39E-02 | 1.70 |
| A0A0C4DH38 | IGHV5-51 | Immunoglobulin heavy variable 5-51 | 99 | 20 | 3.25E-02 | 1.41E-02 | 0.70 |
| P60174 | TPI1 | Triosephosphate isomerase | 63 | 20 | 4.98E-02 | 1.44E-02 | 1.74 |
| P04196 | HRG | Histidine-rich glycoprotein | 110 | 20 | 3.19E-02 | 1.45E-02 | 0.81 |
| Q15942 | ZYX | Zyxin | 58 | 12 | 9.82E-03 | 1.66E-02 | 0.53 |
| P35542 | SAA4 | Serum amyloid A-4 protein | 110 | 20 | 6.30E-03 | 1.68E-02 | 0.75 |
| Q96IY4 | CPB2 | Carboxypeptidase B2 | 110 | 20 | 1.36E-03 | 1.79E-02 | 0.83 |
| Q9UGM5 | FETUB | Fetuin-B | 106 | 19 | 1.42E-03 | 1.86E-02 | 0.73 |
| P25788 | PSMA3 | Proteasome subunit alpha type-3 | 36 | 19 | 1.55E-02 | 2.01E-02 | 1.89 |
| P13639 | EEF2 | Elongation factor 2 | 13 | 18 | 2.32E-02 | 2.12E-02 | 1.81 |
| P00747 | PLG | Plasminogen | 110 | 20 | 1.61E-03 | 2.37E-02 | 0.84 |

| | | | | | | | |
|---|---|---|---|---|---|---|---|
| O75019 | LILRA1 | Leukocyte immunoglobulin-like receptor subfamily A member 1 | 70 | 12 | 1.61E-02 | 2.54E-02 | 0.70 |
| O75874 | IDH1 | Isocitrate dehydrogenase [NADP] cytoplasmic | 61 | 20 | 3.25E-02 | 2.82E-02 | 1.75 |
| P68363 | TUBA1B | Tubulin alpha-1B chain | 67 | 19 | 1.74E-02 | 3.36E-02 | 0.70 |
| P14649 | MYL6B | Myosin light chain 6B | 42 | 10 | 4.21E-02 | 3.53E-02 | 0.63 |
| P52209 | PGD | 6-phosphogluconate dehydrogenase, decarboxylating | 38 | 20 | 2.89E-02 | 3.85E-02 | 1.47 |
| P09871 | C1S | Complement C1s subcomponent | 110 | 20 | 3.17E-03 | 3.91E-02 | 0.86 |
| P51884 | LUM | Lumican | 110 | 20 | 3.34E-03 | 4.09E-02 | 1.25 |
| P04275 | VWF | von Willebrand factor | 110 | 20 | 1.16E-07 | 4.27E-02 | 1.55 |
| A0A0B4J1U3 | IGLV1-36 | Immunoglobulin lambda variable 1-36 | 78 | 17 | 1.33E-02 | 4.36E-02 | 0.87 |
| P42765 | ACAA2 | 3-ketoacyl-CoA thiolase, mitochondrial | 22 | 18 | 1.81E-02 | 4.41E-02 | 1.98 |

**Day 4 - Cluster A vs Cluster B**

| Protein | Gene | Description | Patients cluster A | Patients cluster B | ANOVA $p_{FDR}$-value | Posthoc p-value | Ratio of means B / A |
|---|---|---|---|---|---|---|---|
| P00746 | CFD | Complement factor D | 112 | 82 | 2.83E-04 | 4.48E-06 | 1.67 |
| Q9NZP8 | C1RL | Complement C1r subcomponent-like protein | 112 | 83 | 8.31E-04 | 2.10E-05 | 0.84 |
| P01034 | CST3 | Cystatin-C | 111 | 83 | 1.12E-03 | 3.44E-05 | 1.42 |
| P01717 | IGLV3-25 | Immunoglobulin lambda variable 3-25 | 47 | 17 | 1.09E-03 | 5.82E-05 | 0.33 |
| P61769 | B2M | Beta-2-microglobulin | 112 | 83 | 1.81E-03 | 7.39E-05 | 1.40 |
| P04275 | VWF | von Willebrand factor | 112 | 83 | 1.89E-04 | 2.98E-04 | 1.53 |
| P07478 | PRSS2 | Trypsin-2 | 43 | 48 | 6.57E-03 | 5.46E-04 | 2.22 |
| Q12805 | EFEMP1 | EGF-containing fibulin-like extracellular matrix protein 1 | 105 | 80 | 7.65E-03 | 5.76E-04 | 1.25 |
| P01768 | IGHV3-30 | Immunoglobulin heavy variable 3-30 | 49 | 21 | 7.05E-03 | 8.42E-04 | 0.67 |
| P06744 | GPI | Glucose-6-phosphate isomerase | 21 | 26 | 1.31E-03 | 1.26E-03 | 1.95 |
| P36955 | SERPINF1 | Pigment epithelium-derived factor | 112 | 83 | 1.54E-02 | 1.54E-03 | 1.17 |
| P51884 | LUM | Lumican | 112 | 83 | 7.05E-03 | 1.82E-03 | 1.28 |
| P02741 | CRP | C-reactive protein | 112 | 83 | 2.60E-03 | 2.10E-03 | 1.26 |
| P61626 | LYZ | Lysozyme C | 106 | 83 | 1.68E-02 | 2.44E-03 | 1.26 |
| P08294 | SOD3 | Extracellular superoxide dismutase [Cu-Zn] | 87 | 73 | 2.16E-02 | 2.69E-03 | 1.60 |
| Q9Y279 | VSIG4 | V-set and immunoglobulin domain-containing protein 4 | 75 | 71 | 9.88E-03 | 3.21E-03 | 1.44 |
| P00747 | PLG | Plasminogen | 112 | 83 | 2.83E-04 | 3.85E-03 | 0.90 |
| A0A075B6J9 | IGLV2-18 | Immunoglobulin lambda variable 2-18 | 104 | 67 | 2.14E-02 | 6.50E-03 | 1.44 |
| P10643 | C7 | Complement component C7 | 112 | 83 | 1.54E-02 | 6.50E-03 | 1.20 |
| P04180 | LCAT | Phosphatidylcholine-sterol acyltransferase | 111 | 81 | 1.73E-07 | 6.88E-03 | 0.88 |
| P01024 | C3 | Complement C3 | 112 | 83 | 5.18E-03 | 1.16E-02 | 0.89 |
| P02649 | APOE | Apolipoprotein E | 112 | 83 | 1.54E-02 | 1.17E-02 | 1.20 |

| | | | | | | | |
|---|---|---|---|---|---|---|---|
| P13473 | LAMP2 | Lysosome-associated membrane glycoprotein 2 | 52 | 29 | 1.32E-02 | 1.68E-02 | 0.76 |
| P02760 | AMBP | Protein AMBP | 112 | 83 | 3.33E-02 | 1.69E-02 | 1.19 |
| P09467 | FBP1 | Fructose-1,6-bisphosphatase 1 | 22 | 19 | 3.47E-02 | 1.79E-02 | 2.29 |
| P10909 | CLU | Clusterin | 112 | 83 | 9.30E-04 | 2.04E-02 | 0.93 |
| P02790 | HPX | Hemopexin | 112 | 83 | 2.70E-06 | 2.28E-02 | 0.87 |
| P02774 | GC | Vitamin D-binding protein | 112 | 83 | 7.54E-03 | 3.76E-02 | 0.93 |
| P03951 | F11 | Coagulation factor XI | 110 | 79 | 5.60E-04 | 4.79E-02 | 0.94 |

### Day 4 - Cluster A vs. Cluster C

| Protein | Gene | Description | Patients cluster A | Patients cluster C | ANOVA $p_{FDR}$-value | Posthoc p-value | Ratio of means C / A |
|---|---|---|---|---|---|---|---|
| P04180 | LCAT | Phosphatidylcholine-sterol acyltransferase | 111 | 5 | 1.73E-07 | 3.36E-09 | 0.43 |
| P02790 | HPX | Hemopexin | 112 | 5 | 2.70E-06 | 4.37E-08 | 0.55 |
| P05160 | F13B | Coagulation factor XIII B chain | 111 | 5 | 1.18E-04 | 7.03E-07 | 0.44 |
| P49908 | SELENOP | Selenoprotein P | 112 | 5 | 5.03E-05 | 7.64E-07 | 0.44 |
| P04070 | PROC | Vitamin K-dependent protein C | 107 | 5 | 1.24E-04 | 1.59E-06 | 0.29 |
| P04196 | HRG | Histidine-rich glycoprotein | 112 | 5 | 2.83E-04 | 3.54E-06 | 0.38 |
| P00338 | LDHA | L-lactate dehydrogenase A chain | 99 | 5 | 2.83E-04 | 7.39E-06 | 12.03 |
| P01008 | SERPINC1 | Antithrombin-III | 112 | 5 | 3.74E-04 | 1.51E-05 | 0.56 |
| P02748 | C9 | Complement component C9 | 112 | 5 | 7.06E-04 | 1.54E-05 | 0.55 |
| P04278 | SHBG | Sex hormone-binding globulin | 105 | 5 | 7.44E-04 | 2.38E-05 | 0.44 |
| P12259 | F5 | Coagulation factor V | 112 | 5 | 9.30E-04 | 3.19E-05 | 0.56 |
| P05546 | SERPIND1 | Heparin cofactor 2 | 112 | 5 | 5.60E-04 | 3.74E-05 | 0.47 |
| P03951 | F11 | Coagulation factor XI | 110 | 5 | 5.60E-04 | 4.44E-05 | 0.52 |

| | | | | | | | |
|---|---|---|---|---|---|---|---|
| P11021 | HSPA5 | Endoplasmic reticulum chaperone BiP | 105 | 5 | 1.38E-03 | 5.41E-05 | 3.94 |
| P01042 | KNG1 | Kininogen-1 | 112 | 5 | 1.30E-03 | 7.12E-05 | 0.67 |
| P05062 | ALDOB | Fructose-bisphosphate aldolase B | 89 | 5 | 1.70E-03 | 7.87E-05 | 7.18 |
| P00738 | HP | Haptoglobin | 112 | 5 | 1.89E-03 | 9.77E-05 | 0.31 |
| P00747 | PLG | Plasminogen | 112 | 5 | 2.83E-04 | 1.01E-04 | 0.58 |
| P26927 | MST1 | Hepatocyte growth factor-like protein | 108 | 5 | 1.41E-03 | 2.33E-04 | 0.52 |
| P10909 | CLU | Clusterin | 112 | 5 | 9.30E-04 | 2.39E-04 | 0.59 |
| P02675 | FGB | Fibrinogen beta chain | 112 | 5 | 5.38E-03 | 3.18E-04 | 0.65 |
| P07900 | HSP90AA1 | Heat shock protein HSP 90-alpha | 81 | 5 | 5.18E-03 | 3.76E-04 | 3.62 |
| P04275 | VWF | von Willebrand factor | 112 | 5 | 1.89E-04 | 4.54E-04 | 3.04 |
| P05156 | CFI | Complement factor I | 112 | 5 | 5.18E-03 | 4.78E-04 | 0.57 |
| P04075 | ALDOA | Fructose-bisphosphate aldolase A | 107 | 5 | 5.18E-03 | 5.17E-04 | 2.49 |
| P08185 | SERPINA6 | Corticosteroid-binding globulin | 112 | 5 | 6.74E-03 | 5.25E-04 | 0.57 |
| P37837 | TALDO1 | Transaldolase | 69 | 5 | 7.65E-03 | 6.09E-04 | 3.47 |
| Q14624 | ITIH4 | Inter-alpha-trypsin inhibitor heavy chain H4 | 112 | 5 | 5.94E-03 | 6.37E-04 | 0.66 |
| Q03591 | CFHR1 | Complement factor H-related protein 1 | 112 | 5 | 8.25E-03 | 6.40E-04 | 0.45 |
| P04004 | VTN | Vitronectin | 112 | 5 | 4.51E-03 | 7.93E-04 | 0.59 |
| P20851 | C4BPB | C4b-binding protein beta chain | 108 | 5 | 1.08E-02 | 9.32E-04 | 0.52 |
| P02144 | MB | Myoglobin | 75 | 5 | 6.74E-03 | 1.00E-03 | 46.61 |
| P60709 | ACTB | Actin, cytoplasmic 1 | 112 | 5 | 4.44E-03 | 1.01E-03 | 5.43 |
| P15259 | PGAM2 | Phosphoglycerate mutase 2 | 38 | 5 | 1.27E-02 | 1.24E-03 | 4.39 |
| P0C0S8 | H2AC17 | Histone H2A type 1 | 97 | 5 | 1.15E-02 | 1.33E-03 | 6.87 |
| Q16851 | UGP2 | UTP--glucose-1-phosphate uridylyltransferase | 59 | 5 | 1.55E-02 | 1.61E-03 | 4.13 |
| P04040 | CAT | Catalase | 87 | 5 | 1.44E-02 | 1.64E-03 | 2.17 |
| P29401 | TKT | Transketolase | 80 | 5 | 1.49E-02 | 1.71E-03 | 3.99 |

| | | | | | | | |
|---|---|---|---|---|---|---|---|
| P13796 | LCP1 | Plastin-2 | 112 | 5 | 1.12E-02 | 1.85E-03 | 4.11 |
| P02671 | FGA | Fibrinogen alpha chain | 112 | 5 | 1.57E-02 | 2.22E-03 | 0.72 |
| P22891 | PROZ | Vitamin K-dependent protein Z | 100 | 5 | 2.10E-02 | 2.44E-03 | 0.32 |
| O60814 | H2BC12 | Histone H2B type 1-K | 100 | 5 | 1.75E-02 | 3.39E-03 | 6.06 |
| P08519 | LPA | Apolipoprotein(a) | 103 | 5 | 3.24E-02 | 4.27E-03 | 0.39 |
| P04406 | GAPDH | Glyceraldehyde-3-phosphate dehydrogenase | 80 | 5 | 3.04E-02 | 4.34E-03 | 6.19 |
| P02774 | GC | Vitamin D-binding protein | 112 | 5 | 7.54E-03 | 4.52E-03 | 0.69 |
| P02656 | APOC3 | Apolipoprotein C-III | 112 | 5 | 3.01E-02 | 4.71E-03 | 0.44 |
| P01024 | C3 | Complement C3 | 112 | 5 | 5.18E-03 | 6.20E-03 | 0.65 |
| P02679 | FGG | Fibrinogen gamma chain | 112 | 5 | 4.17E-02 | 6.97E-03 | 0.82 |
| P07357 | C8A | Complement component C8 alpha chain | 112 | 5 | 3.58E-02 | 8.02E-03 | 0.67 |
| P02749 | APOH | Beta-2-glycoprotein 1 | 112 | 5 | 3.47E-02 | 8.74E-03 | 0.61 |
| P21333 | FLNA | Filamin-A | 86 | 5 | 4.98E-02 | 9.48E-03 | 0.32 |
| P13671 | C6 | Complement component C6 | 112 | 5 | 1.67E-02 | 1.18E-02 | 0.69 |
| P29622 | SERPINA4 | Kallistatin | 112 | 5 | 3.80E-02 | 1.26E-02 | 0.58 |
| P02741 | CRP | C-reactive protein | 112 | 5 | 2.60E-03 | 1.26E-02 | 2.30 |
| P01019 | AGT | Angiotensinogen | 112 | 5 | 4.98E-02 | 2.87E-02 | 1.59 |
| Q03154 | ACY1 | Aminoacylase-1 | 27 | 5 | 3.33E-02 | 3.54E-02 | 2.24 |
| P02649 | APOE | Apolipoprotein E | 112 | 5 | 1.54E-02 | 4.85E-02 | 1.55 |

### Day 4 - Cluster B vs. Cluster C

| Protein | Gene | Description | Patients cluster B | Patients cluster C | ANOVA $p_{FDR}$-value | Posthoc p-value | Ratio of means C / B |
|---|---|---|---|---|---|---|---|
| P04180 | LCAT | Phosphatidylcholine-sterol acyltransferase | 81 | 5 | 1.73E-07 | 6.40E-07 | 0.48 |

| | | | | | | | |
|---|---|---|---|---|---|---|---|
| P02790 | HPX | Hemopexin | 83 | 5 | 2.70E-06 | 3.25E-06 | 0.63 |
| P05160 | F13B | Coagulation factor XIII B chain | 81 | 5 | 1.18E-04 | 3.29E-06 | 0.44 |
| P04278 | SHBG | Sex hormone-binding globulin | 77 | 5 | 7.44E-04 | 2.13E-05 | 0.43 |
| P49908 | SELENOP | Selenoprotein P | 83 | 5 | 5.03E-05 | 2.53E-05 | 0.48 |
| P04196 | HRG | Histidine-rich glycoprotein | 83 | 5 | 2.83E-04 | 2.64E-05 | 0.41 |
| P04070 | PROC | Vitamin K-dependent protein C | 74 | 5 | 1.24E-04 | 3.06E-05 | 0.33 |
| P02748 | C9 | Complement component C9 | 83 | 5 | 7.06E-04 | 6.42E-05 | 0.57 |
| P00338 | LDHA | L-lactate dehydrogenase A chain | 67 | 5 | 2.83E-04 | 1.39E-04 | 5.16 |
| P12259 | F5 | Coagulation factor V | 83 | 5 | 9.30E-04 | 2.11E-04 | 0.58 |
| P11021 | HSPA5 | Endoplasmic reticulum chaperone BiP | 80 | 5 | 1.38E-03 | 2.48E-04 | 3.36 |
| P01008 | SERPINC1 | Antithrombin-III | 83 | 5 | 3.74E-04 | 2.66E-04 | 0.60 |
| Q14624 | ITIH4 | Inter-alpha-trypsin inhibitor heavy chain H4 | 83 | 5 | 5.94E-03 | 3.68E-04 | 0.65 |
| P05062 | ALDOB | Fructose-bisphosphate aldolase B | 72 | 5 | 1.70E-03 | 4.57E-04 | 3.85 |
| P00738 | HP | Haptoglobin | 83 | 5 | 1.89E-03 | 4.99E-04 | 0.35 |
| P02675 | FGB | Fibrinogen beta chain | 83 | 5 | 5.38E-03 | 5.49E-04 | 0.62 |
| P08185 | SERPINA6 | Corticosteroid-binding globulin | 83 | 5 | 6.74E-03 | 5.61E-04 | 0.58 |
| P01042 | KNG1 | Kininogen-1 | 83 | 5 | 1.30E-03 | 6.11E-04 | 0.70 |
| P05546 | SERPIND1 | Heparin cofactor 2 | 83 | 5 | 5.60E-04 | 7.06E-04 | 0.52 |
| P03951 | F11 | Coagulation factor XI | 79 | 5 | 5.60E-04 | 9.99E-04 | 0.55 |
| Q03591 | CFHR1 | Complement factor H-related protein 1 | 81 | 5 | 8.25E-03 | 1.47E-03 | 0.48 |
| P02671 | FGA | Fibrinogen alpha chain | 83 | 5 | 1.57E-02 | 1.72E-03 | 0.68 |
| P20851 | C4BPB | C4b-binding protein beta chain | 76 | 5 | 1.08E-02 | 1.81E-03 | 0.56 |
| P07900 | HSP90AA1 | Heat shock protein HSP 90-alpha | 69 | 5 | 5.18E-03 | 2.23E-03 | 2.36 |
| P37837 | TALDO1 | Transaldolase | 66 | 5 | 7.65E-03 | 2.28E-03 | 2.73 |
| P05156 | CFI | Complement factor I | 83 | 5 | 5.18E-03 | 2.94E-03 | 0.59 |

| | | | | | | | |
|---|---|---|---|---|---|---|---|
| P04075 | ALDOA | Fructose-bisphosphate aldolase A | 79 | 5 | 5.18E-03 | 3.47E-03 | 2.13 |
| P26927 | MST1 | Hepatocyte growth factor-like protein | 72 | 5 | 1.41E-03 | 4.05E-03 | 0.58 |
| P02656 | APOC3 | Apolipoprotein C-III | 83 | 5 | 3.01E-02 | 4.11E-03 | 0.39 |
| P00747 | PLG | Plasminogen | 83 | 5 | 2.83E-04 | 4.75E-03 | 0.64 |
| P02749 | APOH | Beta-2-glycoprotein 1 | 83 | 5 | 3.47E-02 | 4.93E-03 | 0.59 |
| P04406 | GAPDH | Glyceraldehyde-3-phosphate dehydrogenase | 68 | 5 | 3.04E-02 | 5.04E-03 | 5.11 |
| P07357 | C8A | Complement component C8 alpha chain | 83 | 5 | 3.58E-02 | 5.15E-03 | 0.65 |
| Q03154 | ACY1 | Aminoacylase-1 | 37 | 5 | 3.33E-02 | 5.17E-03 | 2.92 |
| Q16851 | UGP2 | UTP--glucose-1-phosphate uridylyltransferase | 39 | 5 | 1.55E-02 | 5.17E-03 | 2.38 |
| P10909 | CLU | Clusterin | 83 | 5 | 9.30E-04 | 5.42E-03 | 0.63 |
| P22891 | PROZ | Vitamin K-dependent protein Z | 60 | 5 | 2.10E-02 | 5.50E-03 | 0.35 |
| P0C0S8 | H2AC17 | Histone H2A type 1 | 71 | 5 | 1.15E-02 | 5.87E-03 | 2.79 |
| P08519 | LPA | Apolipoprotein(a) | 76 | 5 | 3.24E-02 | 6.55E-03 | 0.42 |
| P04040 | CAT | Catalase | 64 | 5 | 1.44E-02 | 6.84E-03 | 1.69 |
| P02679 | FGG | Fibrinogen gamma chain | 83 | 5 | 4.17E-02 | 7.32E-03 | 0.76 |
| P15259 | PGAM2 | Phosphoglycerate mutase 2 | 38 | 5 | 1.27E-02 | 7.52E-03 | 3.59 |
| P04004 | VTN | Vitronectin | 83 | 5 | 4.51E-03 | 7.84E-03 | 0.63 |
| P29401 | TKT | Transketolase | 54 | 5 | 1.49E-02 | 7.87E-03 | 2.82 |
| P21333 | FLNA | Filamin-A | 71 | 5 | 4.98E-02 | 8.84E-03 | 0.32 |
| P13796 | LCP1 | Plastin-2 | 82 | 5 | 1.12E-02 | 1.01E-02 | 3.39 |
| P02144 | MB | Myoglobin | 61 | 5 | 6.74E-03 | 1.04E-02 | 24.02 |
| P60709 | ACTB | Actin, cytoplasmic 1 | 83 | 5 | 4.44E-03 | 1.12E-02 | 3.38 |
| O60814 | H2BC12 | Histone H2B type 1-K | 72 | 5 | 1.75E-02 | 1.74E-02 | 2.53 |
| P04275 | VWF | von Willebrand factor | 83 | 5 | 1.89E-04 | 2.84E-02 | 1.98 |
| P02774 | GC | Vitamin D-binding protein | 83 | 5 | 7.54E-03 | 4.54E-02 | 0.75 |

**Supplementary Table 8** Feature importance ranks for random forest classifier. Classifier was trained 100 times in a Monte Carlo cross-validation and the features were interpreted using Shapley Additive Explanations (SHAP). Table shows median rank of one random iteration.

| | Feature | Median rank |
|---|---|---|
| 1 | ALT_MAX | 2.0 |
| 2 | AST_MAX | 2.0 |
| 3 | INR_MAX | 3.0 |
| 4 | BE_MEDIAN | 4.0 |
| 5 | BP_dia_MEDIAN | 4.0 |
| 6 | APTT_MAX | 7.0 |
| 7 | BP_sys_MEDIAN | 8.0 |
| 8 | pH_MIN | 8.0 |
| 9 | PCT_MAX | 9.0 |
| 10 | CREA_MAX | 10.0 |
| 11 | NA_MIN | 12.0 |
| 12 | K_MAX | 13.0 |
| 13 | PROT_total_MIN | 13.0 |
| 14 | CA_ionized_MIN | 16.0 |
| 15 | WBC_MIN | 16.0 |
| 16 | FIBR_MIN | 18.0 |
| 17 | SpO2_MIN | 18.0 |
| 18 | UREA_N_MAX | 18.0 |
| 19 | ALP_MAX | 19.0 |
| 20 | BR_total_MAX | 20.0 |
| 21 | TC_MAX | 21.0 |
| 22 | HGB_MIN | 22.0 |
| 23 | PLT_MIN | 24.0 |
| 24 | GGT_MAX | 24.0 |
| 25 | PH_MIN | 24.0 |
| 26 | CK_MAX | 26.0 |
| 27 | FiO2_MAX | 26.0 |
| 28 | pCO2_MAX | 27.0 |
| 29 | HR_MEDIAN | 27.0 |
| 30 | pO2_MIN | 29.0 |
| 31 | CRP_MAX | 30.0 |
| 32 | LIP_MAX | 31.0 |
| 33 | TEMP_MAX | 31.0 |
| 34 | TG_MAX | 32.0 |

**Supplementary Table 9:** Machine learning model performance metrics.

| | Test Set | | Training Set | |
|---|---|---|---|---|
| **Metric** | **Mean** | **SD** | **Mean** | **SD** |
| **Accuracy** | 0.833 | 0.0342 | 0.953 | 0.011 |
| **AUROC** | 0.950 | 0.016 | 0.995 | 0.002 |
| **F1 Score** | 0.821 | 0.044 | 0.947 | 0.012 |
| **MCC** | 0.703 | 0.060 | 0.915 | 0.019 |
| **Precision** | 0.813 | 0.050 | 0.933 | 0.017 |
| **Recall** | 0.839 | 0.043 | 0.965 | 0.009 |

### Supplementary Figures

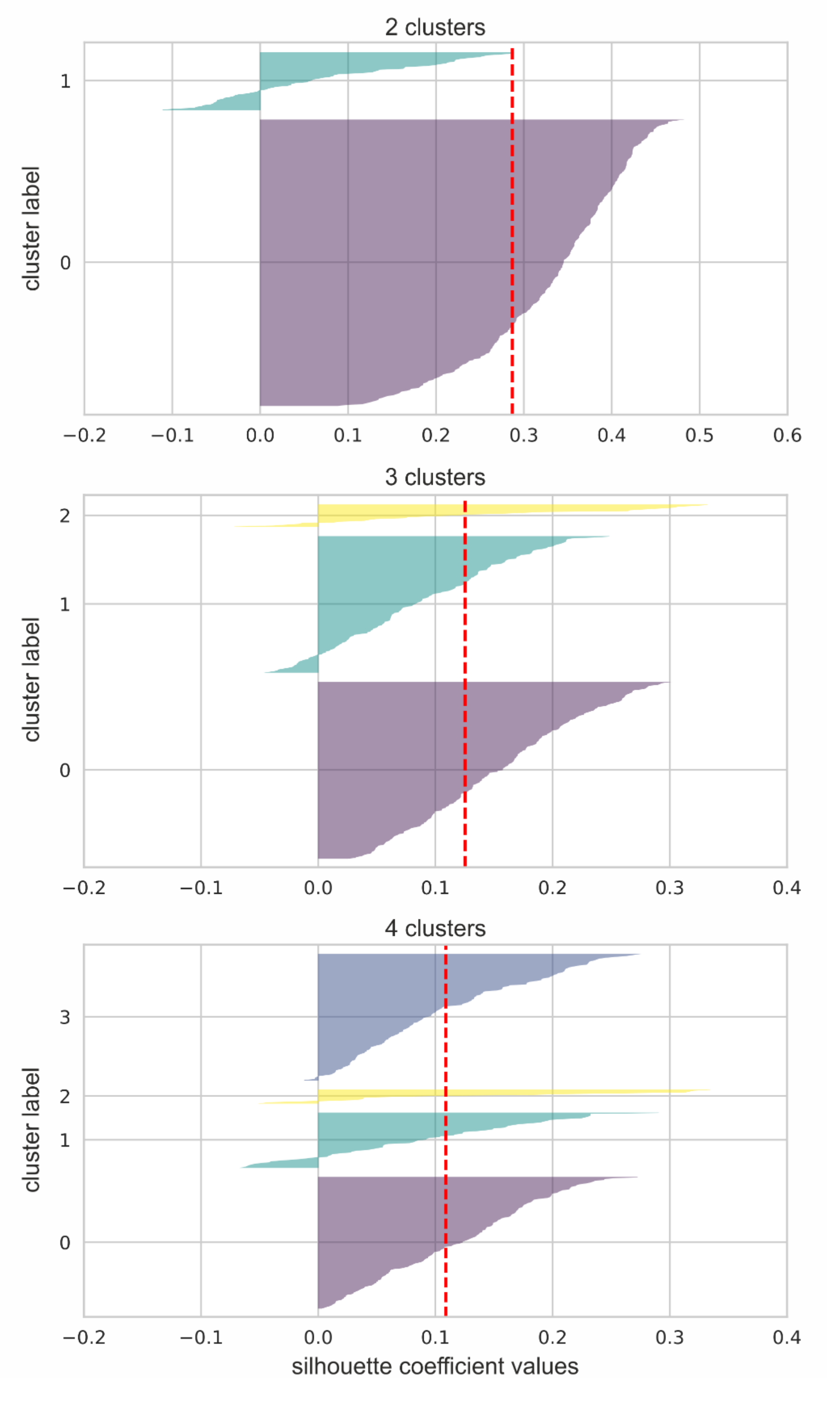


**Supplementary Figure 1:** Silhouette curves that were used for determination of the optimal number of clusters. Red dotted line representing the mean silhouette coefficient. In the case of two clusters, there was a large deviation between the silhouette coefficients (0.29 ± 0.13) and one cluster was clearly smaller than the other. Three clusters show the highest mean silhouette coefficient (0.13 ± 0.08). Using four clusters, the mean silhouette coefficient was slightly lower than with three clusters (0.09 ± 0.08) and the number of points in the cluster decreased.

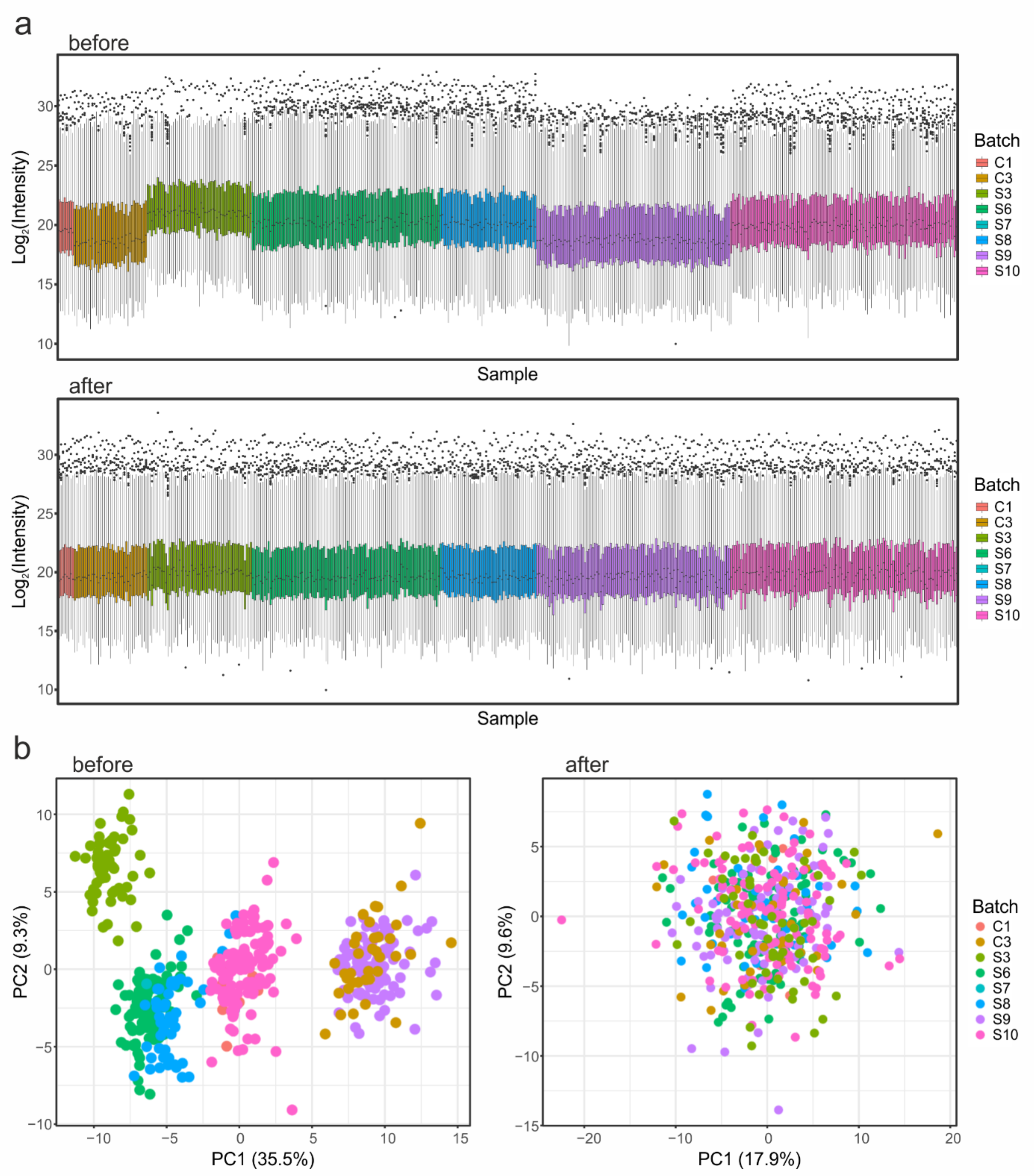


**Supplementary Figure 2:** Quality control of proteomics batch normalization. **A** Boxplot representation of protein intensities before and after normalization. Boxes indicate the 25% - 75% interquartile range (IQR) with the median displayed as horizontal line. Whiskers extend to 1.5 x IQR. Outliers displayed as individual data points. Each box corresponds to a sample, colors representing the respective batches. **B** Principal component analysis (PCA) plots of mass spectrometry data before and after batch normalization. Each data point corresponds to a sample, colors representing the respective batches.

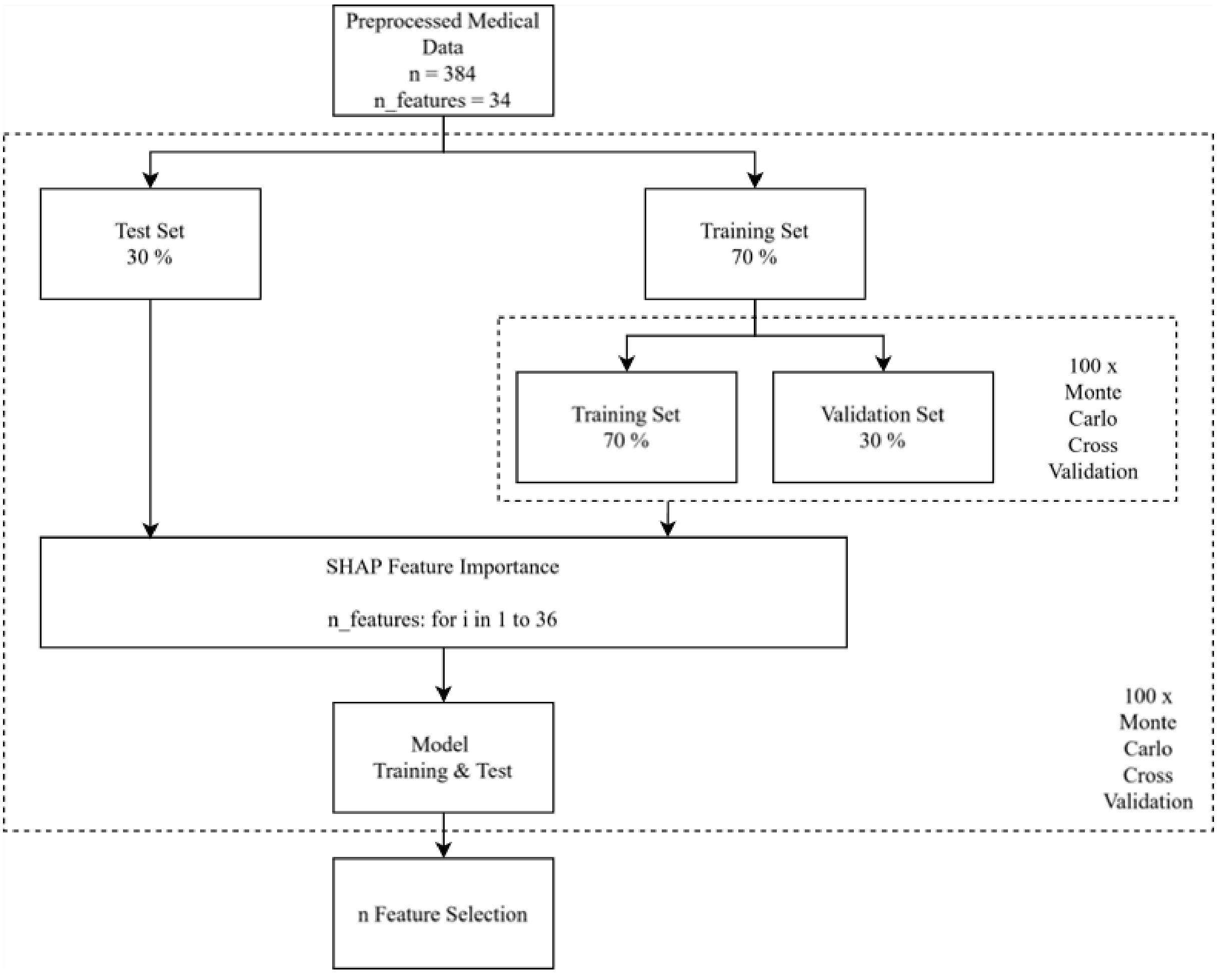


**Supplementary Figure 3**: Framework for Machine Learning and Feature Selection.

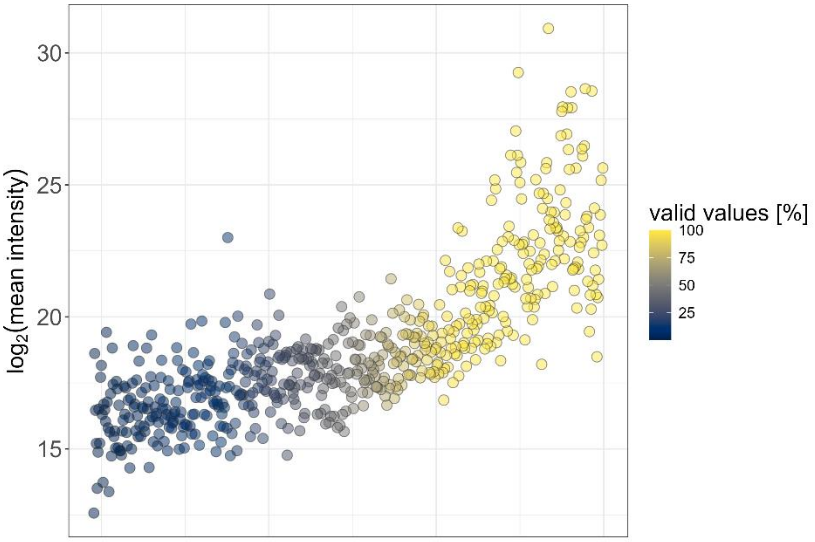


**Supplementary Figure 4:** Illustration of the quantified plasma proteome. Proteins are plotted against their relative intensity. Color represents the percentage of observations in % (100 % correspond to detection of the respective protein in all patients). The figure illustrates the relation of protein abundance and detectability. Approximately half of the quantified proteins (309 of 609) were quantified with at least 50 % valid values.

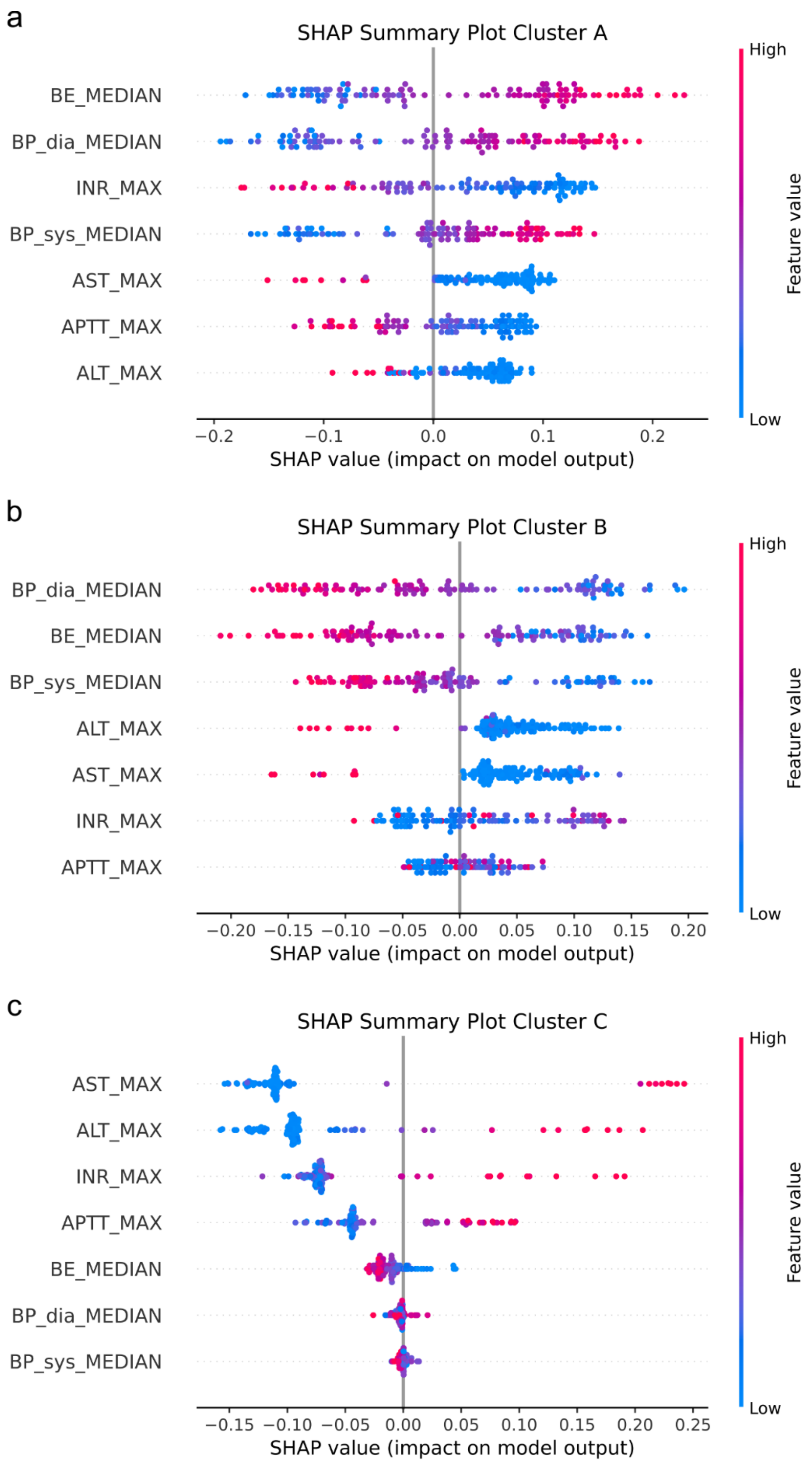


**Supplementary Figure 5:** Exemplary SHAP summary plots of cluster **A**, cluster **B** and cluster **C** showing the impact of each feature on the model's prediction for one random training-test split performed within the MCCV. Colors indicate feature values (blue = low, pink = high).

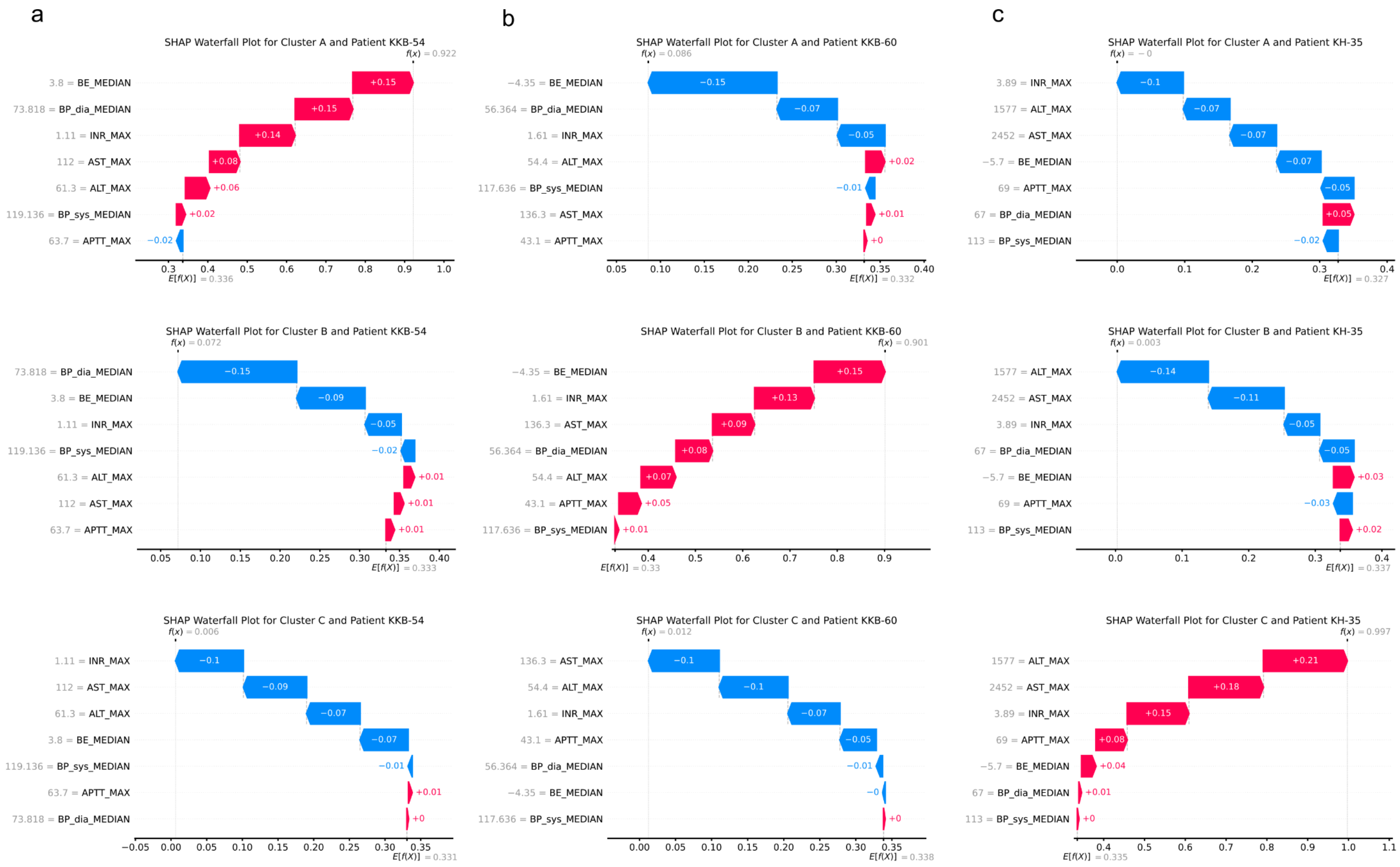

a
b
c
SHAP Waterfall Plot for Cluster A and Patient KKB-54
SHAP Waterfall Plot for Cluster A and Patient KKB-60
SHAP Waterfall Plot for Cluster A and Patient KH-35
SHAP Waterfall Plot for Cluster B and Patient KKB-54
SHAP Waterfall Plot for Cluster B and Patient KKB-60
SHAP Waterfall Plot for Cluster B and Patient KH-35
SHAP Waterfall Plot for Cluster C and Patient KKB-54
SHAP Waterfall Plot for Cluster C and Patient KKB-60
SHAP Waterfall Plot for Cluster C and Patient KH-35

**Supplementary Figure 6:** Exemplary SHAP waterfall plots illustrating feature importance for a representative patient from cluster **A**, cluster **B**, and cluster **C**, derived from random training-test splits in which the patient was included in the test set. The first row corresponds to Cluster A, the second to Cluster B, and the third to Cluster C. Red features indicate a positive contribution for the patient to be classified into the respective Cluster, while blue features indicate a negative contribution. The length of the arrow indicates how much the model output is moved from the prior expectation value $E[f(x)] = 0.33$. The sum of all contributions results in the final value $f(x)$, which represents how confident the model classified the patient into the respective cluster. The original measurements are displayed next to the feature names.
